# Impact of Digital Health Interventions on Health Literacy: A Systematic Review with Quality Appraisal

**DOI:** 10.1101/2025.02.27.25323025

**Authors:** Francesco Andrea Causio, Stefano Gandolfi, Jasleen Kaur, Berna Sert, Maya Fakhfakh, Luigi De Angelis, Marcello Di Pumpo, Giacomo Diedenhofen, Alessandro Berionni, Caterina Rizzo, Fidelia Cascini, Plinio Morita, Timothy Mackey

## Abstract

In the digital era, health literacy is crucial for informed health decisions and improved outcomes. This systematic review examines the effectiveness of digital health interventions (DHIs) in improving health literacy, as defined by the WHO. We included studies (cross-sectional, surveys, and case reports), focusing on interventions like mobile health apps, online platforms, and telehealth services. Our search, adhering to PRISMA guidelines, spanned databases PubMed, IEEE, and ACM, covering publications from 2013 to 2024. From 1.029 initial articles, 39 met our inclusion criteria. Our findings highlight that DHIs, including multimedia tools and remote sessions, have been reported to improve health literacy across diverse populations. However, the impact varies due to the digital divide, influenced by factors like age and socioeconomic status. This review categorizes the included studies by digital intervention type, including: 10.3 % on mobile apps (n = 4), 30.8 % on websites and online platforms (n = 12), 5.1 % on multimedia tools (n = 2), 15.4 % on telehealth and mHealth (n = 6), 2.6 % on electronic health records and patient portals (n = 1), 17.9 % conference proceedings (n = 7), and 17.9 % review studies (n = 7). This study highlights the complex nature of DHIs designed to enhance health literacy and engagement, aiming to reduce health disparities and ensure equitable access to healthcare benefits regardless of socioeconomic background or digital literacy. It underscores the importance of user-centered design, cultural sensitivity, and ongoing support for maximizing the effectiveness of DHIs. Incorporating theoretical frameworks is shown to boost engagement and promote behavioral change, particularly by addressing intrinsic motivations and cultural factors. The findings emphasize the necessity of sustained strategies, such as gamification, to maintain improvements in health literacy, and advocate for standardized evaluation methods to guide policy and advance the global transition to digital-first healthcare.

**Author Summary:** How effective are digital health interventions in improving health literacy? This review of 39 studies found that digital health interventions, including mobile apps, websites, and telehealth, are reported to improve health literacy. Effectiveness of these tools depended on age, digital skills, and socioeconomic factors, with user-centered design, cultural competence, and sustained engagement being key to success. In summary, digital health interventions can improve health literacy but require cultural relevance and sustained engagement for success. The three main takeaways from this collaborative work are:

1. Well-designed digital health interventions can improve health literacy.
2. The digital divide limits access, especially for vulnerable groups.
3. Long-term success requires support, user focus, and cultural relevance.

## 1. Introduction

With approximately 5.5 billion people (more than 2/3rds of the world’s population) currently using the internet, digital tools are at the forefront of our ability to access information and services, including for health. In this increasing digital age, promoting health literacy is a key component in shaping individuals’ health outcomes [1]. Health literacy is defined by the World Health Organization (WHO) as the ability of individuals to gain access to, understand and use information in ways which promote and maintain good health for themselves, their families and their communities [2].

The range of interventions designed to enhance health literacy is broad and varied. Those include small group sessions, digital communications such as text or social media messages, animated movies, multimedia learning tools, applications, and personalized education [3,4]. A subset of these approaches utilizes digital health tools, referred to as digital health or eHealth interventions. According to the WHO classification framework of digital health interventions (DHIs) [5], DHIs encompass an array of technologies and practices aimed at leveraging digital advancements to enhance health outcomes, including artificial intelligence, big data, and telemedicine, among others [6,7].

DHIs have shown promise in promoting healthy behaviors, managing long-term conditions, and facilitating access to treatments, thereby underscoring their significant potential in advancing shared public health goals of improving access to healthcare services, delivering targeted interventions to specific populations. These interventions have been used to encourage behaviors such as smoking cessation, healthy eating, promoting physical activity, and to improve outcomes for individuals with chronic conditions like cardiovascular disease and diabetes [3,8]. Moreover, DHIs offer the possibility of remote access to treatments, such as computerized cognitive behavioral therapy, which is particularly relevant in the context of mental health [9,10].

Nonetheless, the adoption and effectiveness of DHIs is nuanced by the digital divide, a phenomenon that delineates the gap between those who have access to and can utilize information technology and those who do not. This divide is associated with factors such as age, education, income, and urban versus rural living, and is intricately linked to e-health literacy levels [11]. Hence, it is equally crucial for DHIs and associated policy to address the digital divide, which can better ensure that the benefits of digital health interventions are equitably distributed, thereby contributing to the reduction of health disparities.

Previous review articles have investigated the use of digital interventions to improve health literacy across different populations (e.g., older adults, migrant populations) and health domains (e.g., mental health, chronic diseases) investigating computer-based and internet-delivered interventions, often utilizing interactive multimedia content that were generally reported as effective at improving health knowledge, attitudes, and in some cases, behaviors. However, results were mixed for some outcomes [12–17]. While many studies reported improvements in health literacy measures, not all interventions led to significant changes in clinical outcomes or long-term behavior change [12]. While the primary objective of DHIs is to enhance health outcomes, their role in improving health literacy depends on how they are designed and implemented. Some DHIs explicitly integrate educational content that helps users understand health information and make informed decisions, fostering health literacy [3]. As DHIs rapidly evolve due to the introduction of new technologies, there is a need to reassess the evidence supporting the use of DHIs to address health literacy goals.

Our study conducts an interdisciplinary systematic review of the implementation of digital interventions to improve health literacy [18]. By providing an up-to-date description of existing technologies, we aim at providing clinicians and policymakers with the best evidence and description of tools to improve their understanding of such technologies and their impact.

## 2. Materials and Methods

We conducted a multilingual literature search of three major databases: PubMed, IEEE, and ACM. The search strategy was designed to capture studies related to digital interventions for health literacy published between 2013 and 2024 in research indexed for health and biomedical sciences (PubMed), engineering (IEEE), and computer science (ACM). We chose a 10-year literature review period as the aims of this review focused on relatively new or innovative technologies. We used the following search strings, adapted for each database. Specifically, this review updated prior evidence on the topic, including a 2014 systematic review by *Jacobs et al* [12].

For PubMed: ("health literac*"[Title/Abstract] OR "medical knowledge"[Title/Abstract] OR "health knowledge"[Title/Abstract] OR "health literacy"[MeSH Terms] OR "health competence"[Title/Abstract] OR "information literacy"[MeSH Terms]) AND ("digital literac*"[Title/Abstract] OR "Computer literacy"[MeSH Terms] OR "Digital Divide"[MeSH Terms] OR "eHealth literacy"[Title/Abstract] OR "mHealth literacy"[Title/Abstract] OR "Media literacy"[Title/Abstract] OR "electronic health literacy" [Title/Abstract]) AND (2013:2023[pdat]).

For IEEE and ACM: ("health literacy" OR "medical knowledge" OR "health knowledge" OR "health competence" OR "information literacy") AND ("digital literacy" OR "computer literacy" OR "digital divide" OR "eHealth literacy" OR "mHealth literacy" OR "media literacy" OR "electronic health literacy").

### 2.1. Inclusion Criteria

We included original research, cross-sectional studies, surveys, case reports, and other types of primary studies, as well as reviews, commentaries, and observational studies, when they included a description of a digital intervention aimed at increasing health literacy. Articles in English, French, German, and Italian published from 2013 to 2024 were considered eligible. For interventional studies, we used the following PICOT framework:

- Population (P): Any population.
- Intervention (I): Any technological intervention in digital health as defined by the World Health Organization [5]. Digital health, as mentioned within the introduction, is defined as the ability of individuals to gain access to, understand and use information in ways which promote and maintain good health for themselves, their families and their communities.
- Control (C): Not defined.
- Outcome (O): Any outcomes related to health literacy as defined by the Center for Disease Control and Prevention (CDC) [19,20].
- Time (T): Papers/proceedings published after 2013.

We excluded studies focusing on digital health interventions influencing literacy other than health/eHealth or digital outcomes not related to health. Additionally, we excluded digital interventions that were not included in the WHO definition of digital health. Table 1 summarizes the inclusion criteria.

**Table 1:** Inclusion and exclusion criteria.

| **Table 1: Inclusion and exclusion criteria** |  |
| --- | --- |
| **Inclusion Criteria** | **Details** |
| Population | Any population |
| Intervention | Any technological intervention in digital health (WHO definition), but also pertinent non interventional studies are to be included |
| Outcome | Any outcomes related to health literacy (CDC definition). |
| Publication Interval | January 2013-January 2024 |
| Language | Studies in English, French, German, and Italian |
| Study type | Original research, cross-sectional studies, surveys, case reports, and other types of primary studies, reviews, commentaries, and observational studies. |

### 2.2. Study Selection

The study selection process consisted of three phases:

1. Initial search: We identified a total of 1029 articles across the three databases (PubMed, n = 682, IEEE, n = 32, ACM, n = 315) after duplicate removal.
2. Title and abstract screening: Five pairs of reviewers independently screened the titles and abstracts. This screening was conducted in double-blind. Any conflicts were resolved by a third reviewer (FAC). At the end, 124 papers were included for full-text screening.
3. Full-text screening: A total of six authors were randomly assigned 20 or 21 manuscripts each to evaluate for inclusion or exclusion. In cases of uncertainty, a second reviewer (FAC) made the final determination. By the end of this phase, 46 articles were included. During the data extraction phase, conducted using an Excel table (see Table 4), 7 additional papers were excluded for not being exclusively focused on health literacy, leaving 39 articles for manuscript preparation.

### 2.3. Data Extraction & Synthesis

We created a standardized data extraction form in Google Sheets, encompassing over 32 elements for each included paper (see Table 2). For primary studies, seven reviewers extracted data from approximately seven articles each. For systematic reviews, two dedicated reviewers extracted and reported the data. The data extraction process included separating reviews and non-primary articles from primary studies. Then, we conducted a narrative synthesis of the extracted data, supplemented by comparative analysis. Studies were categorized by the type of technology employed to promote health literacy and the methodological design used, including systematic reviews and quantitative versus qualitative approaches.

**Figure 1.**
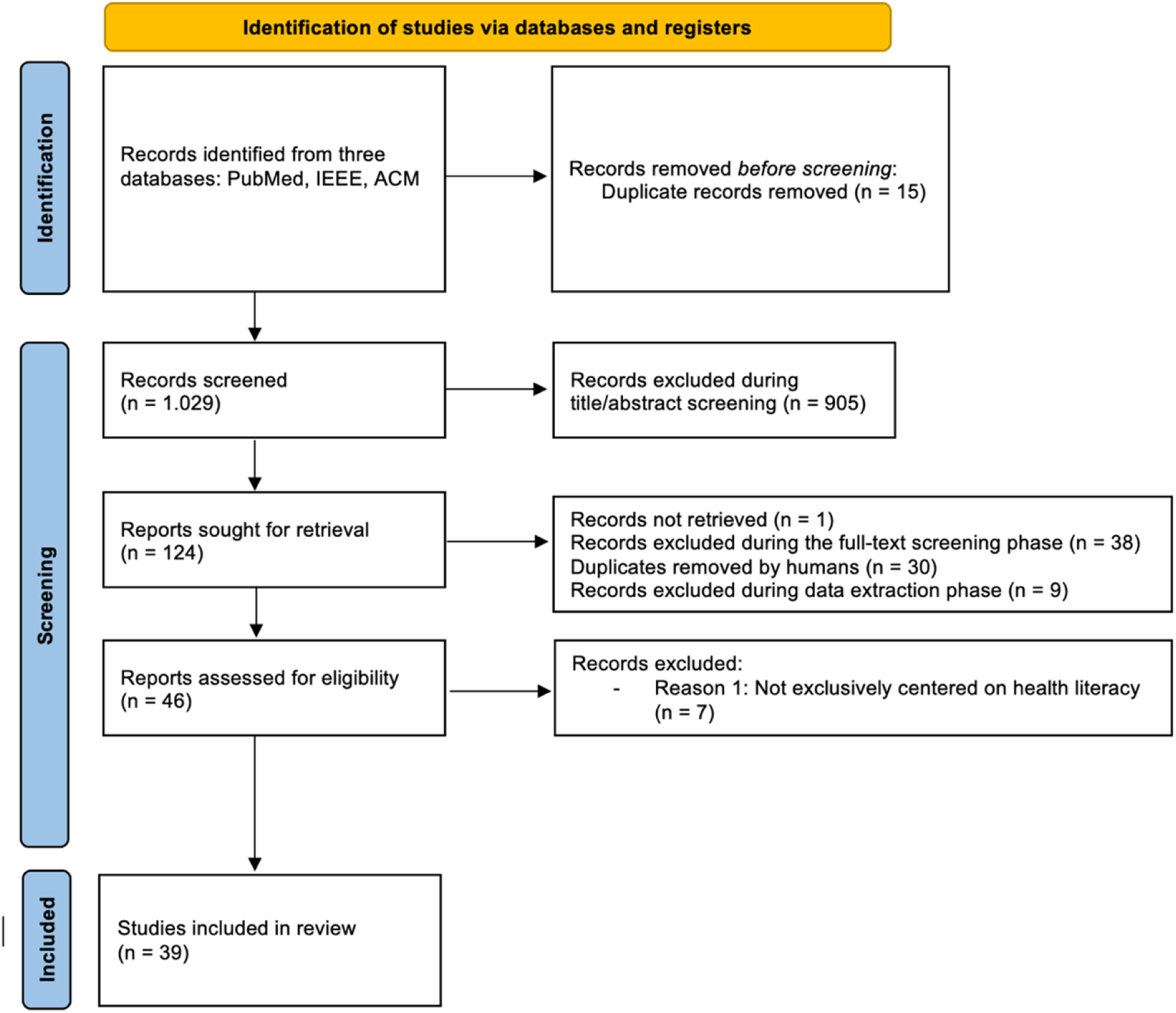
PRISMA flow diagram

**Table 2:** Data dictionary for data extraction.

| n° | VARIABLES | EXPLANATION |
| --- | --- | --- |
| 1 | Study ID | Unique identifier for the study |
| 2 | Authors | Manuscript authors (Names of individuals who contributed to the work). If more than three, report the ones remaining as "et al." |
| 3 | Title | Manuscript title (Name of the study or paper). |
| 4 | Year of publication | When the study or paper was published. Different from the year of the intervention and the year where the manuscript was submitted. |
| 5 | Type of paper | Classification of the paper (conference proceedings, commentary, review, original paper. For the latter, specify the study design as stated in the methods. The most frequent ones are observational studies and interventional studies, including trials). |
| 6 | Journal | Journal in which the study appears |
| 7 | Aim of the study or target health problem and solution | Primary focus or objective of the research (likely declared in the introduction or objective sections). This should include the addressed problem and the identified solution to face it. |
| 8 | Participants | Individuals or groups participating in the study should be the ones who use/are targeted by the digital intervention or tool and upon which the improvement in health literacy is measured. |
| 9 | Age | The age of participants, as detailed in the methods (e.g., age range, average age, and so on) |
| 10 | Gender | Gender distribution of participants |
| 11 | Ethnicity | Ethnic background of participants |
| 12 | Addressed condition(s) | Health issues or conditions being investigated |
| 13 | Total number of participants | Number of individuals participating in the study. If the intervention has more steps, highlight any issues with participants dropping out of the study in the "Retention challenges" column |
| 14 | Method of recruitment of participants | How participants were selected or invited |
| 15 | Study duration | Length of time the study was conducted. If the intervention is shorter than the overall study duration, specify it (e.g., because of a subsequent follow-up phase) |
| 16 | Intervention | Detailed explanation of the treatment or intervention applied |
| 17 | Digital device/technology category | Brief description of the technology used (e.g., mobile app, web portal) |
| 18 | Geographical location(s) of the intervention | Where the study took place |
| 19 | Intervention setting | The context in which the intervention was applied (e.g., urban/suburban/rural; hospital/health district/patients' homes) |
| 20 | Comparator | What the intervention was compared against, if applicable |
| 21 | Outcome(s) | Results of effects observed from the intervention |
| 22 | Outcome(s) measure(s) | How outcomes were quantified or measured |
| 23 | Results | Summary of the findings |
| 24 | Efficacy or conclusion of the intervention(s) | The overall effectiveness of the intervention (as stated in the paper, e.g., positive or negative effect/no effect or no statistically significant difference observed) |
| 25 | Health Guidelines | Whether the study was tailored to existing health guidelines (if any) |
| 26 | eHealth/digital Literacy component | Digital or eHealth literacy component (if any) |
| 27 | eHealth/digital literacy assessment scale used | The specific scale used to measure eHealth/digital literacy (if any) |
| 28 | Cognitive Theory used | Theory applied in the study (if any, eg: Technology Acceptance Model (TAM) or the Unified Theory of Acceptance and Use of Technology (UTAUT)) |
| 29 | Economic information | Cost of resource information related to the study (if any) |
| 30 | Health equity component(s) | Aspects of the study addressing health equity (if any) |
| 31 | Misinformation component(s) | Elements of the study dealing with misinformation (if any) |
| 32 | Retention Challenges | Difficulties encountered in ensuring that participants remain engaged and complete all phases of the study (e.g., usability, acceptability, time burden or complexity of the intervention) |
| 33 | Technological challenges | Any reported technical issues or challenges faced by participants during the study (e.g., connectivity/internet issues, hardware, other) |
| 34 | Notes | Additional remarks or observations |

**Table 3:** Quality Rating Scheme of Oxford Levels of Evidence.

2.4. Quality Assessment The Oxford Levels of Evidence tool was adopted for the quality assessment of the study (see Table 4) [21].
| Table 3: Quality Rating Scheme of Oxford Levels of Evidence |  |
| --- | --- |
| Level 1 | Properly powered and conducted randomized clinical trial; systematic review with meta-analysis |
| Level 2 | Well-designed controlled trial without randomization; prospective comparative cohort trial |
| Level 3 | Case-control studies; retrospective cohort study |
| Level 4 | Case series with or without intervention; cross-sectional study |
| Level 5 | Opinion of respected authorities; case reports |

### 2.4. Quality Assessment

The Oxford Levels of Evidence tool was adopted for the quality assessment of the study (see Table 4) [21].

**Table 4:**
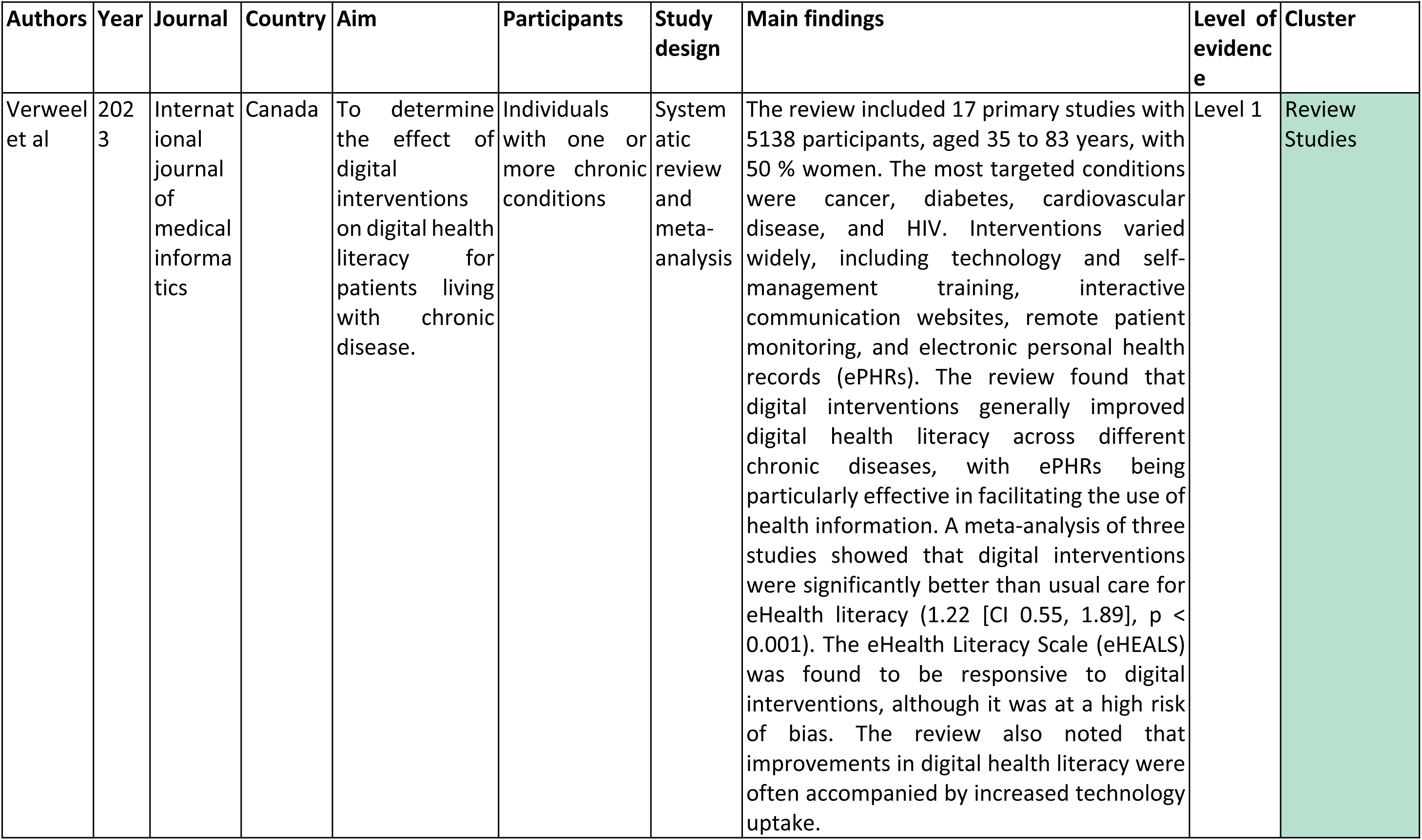
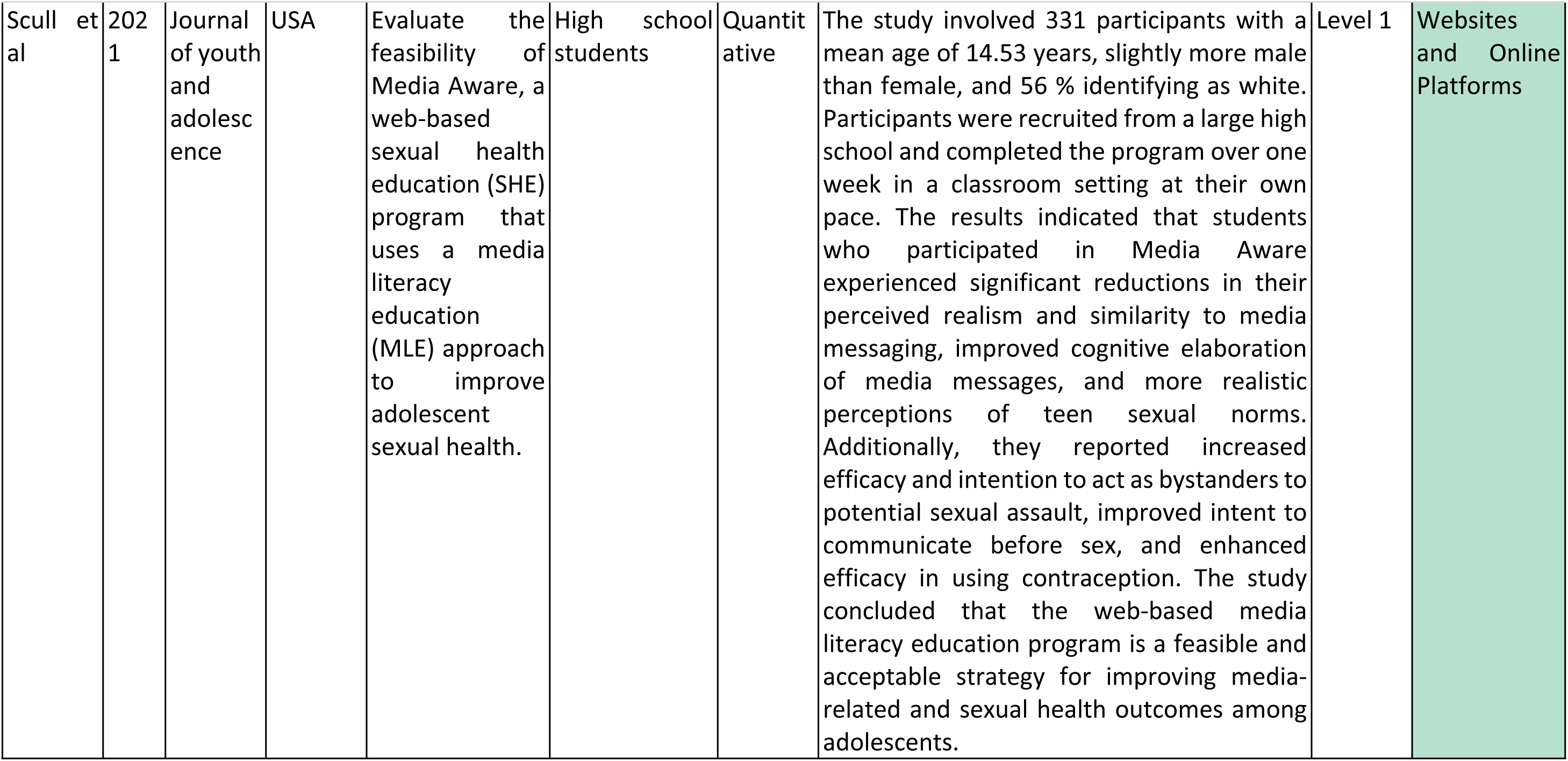
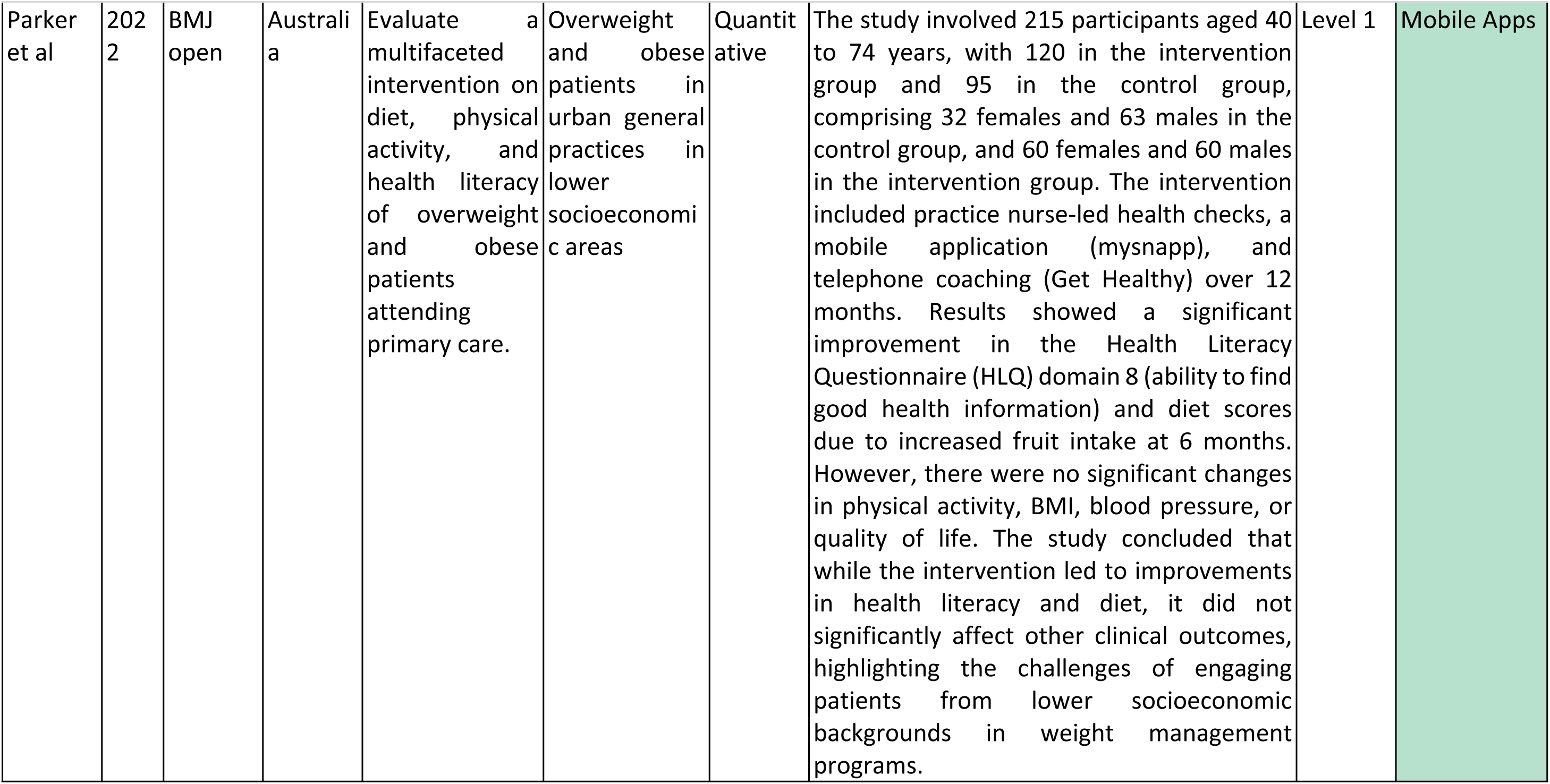
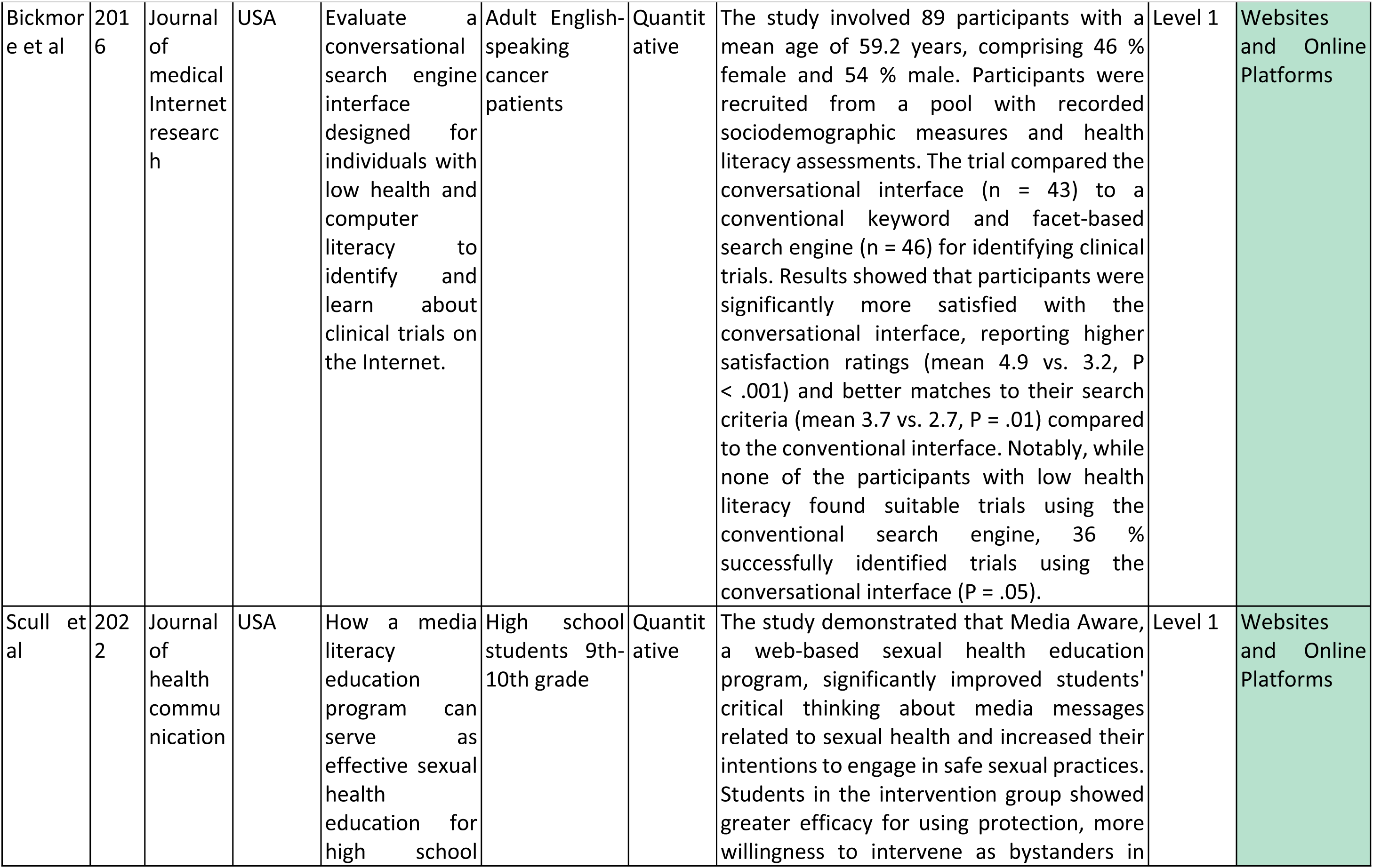
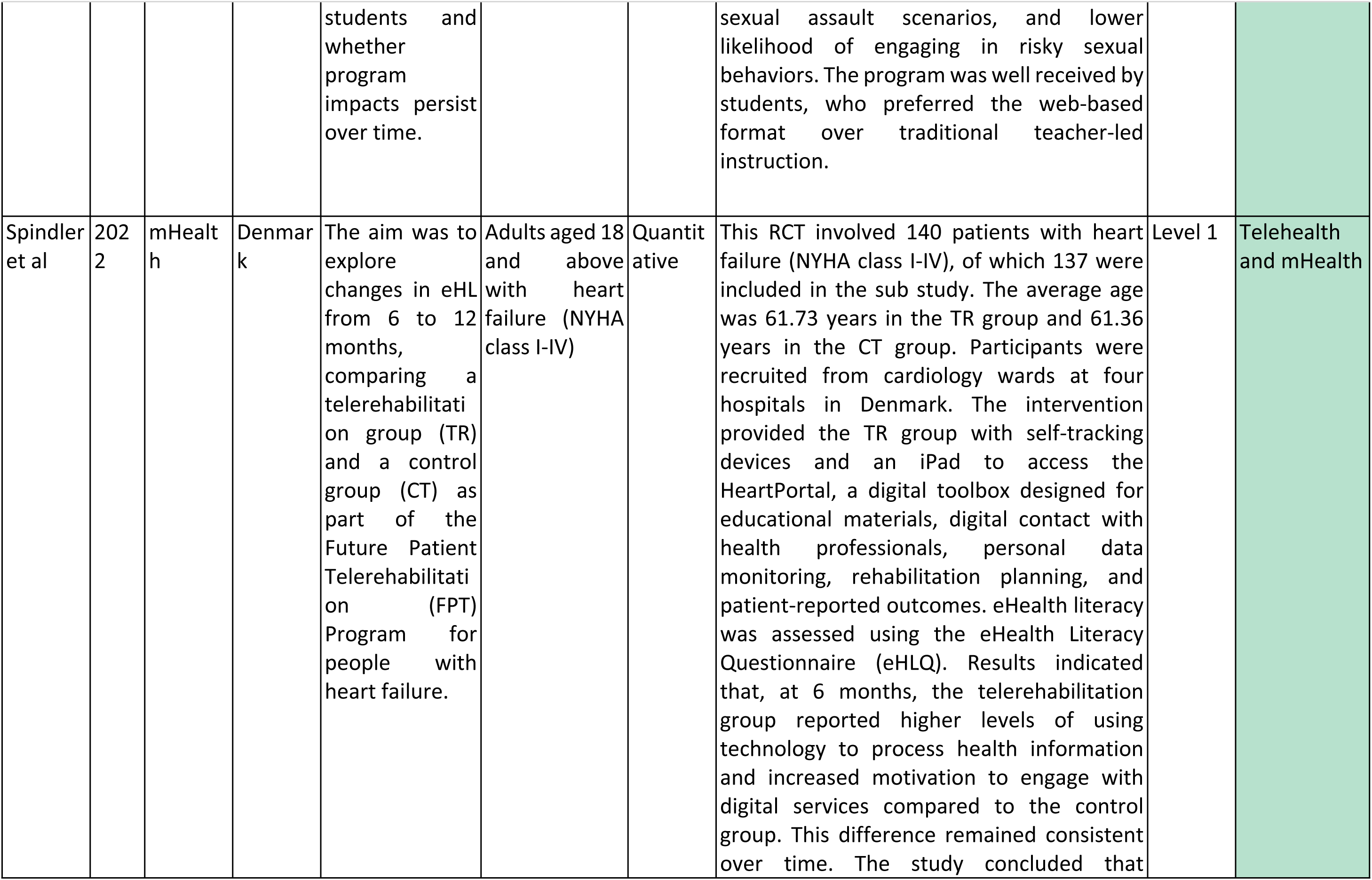
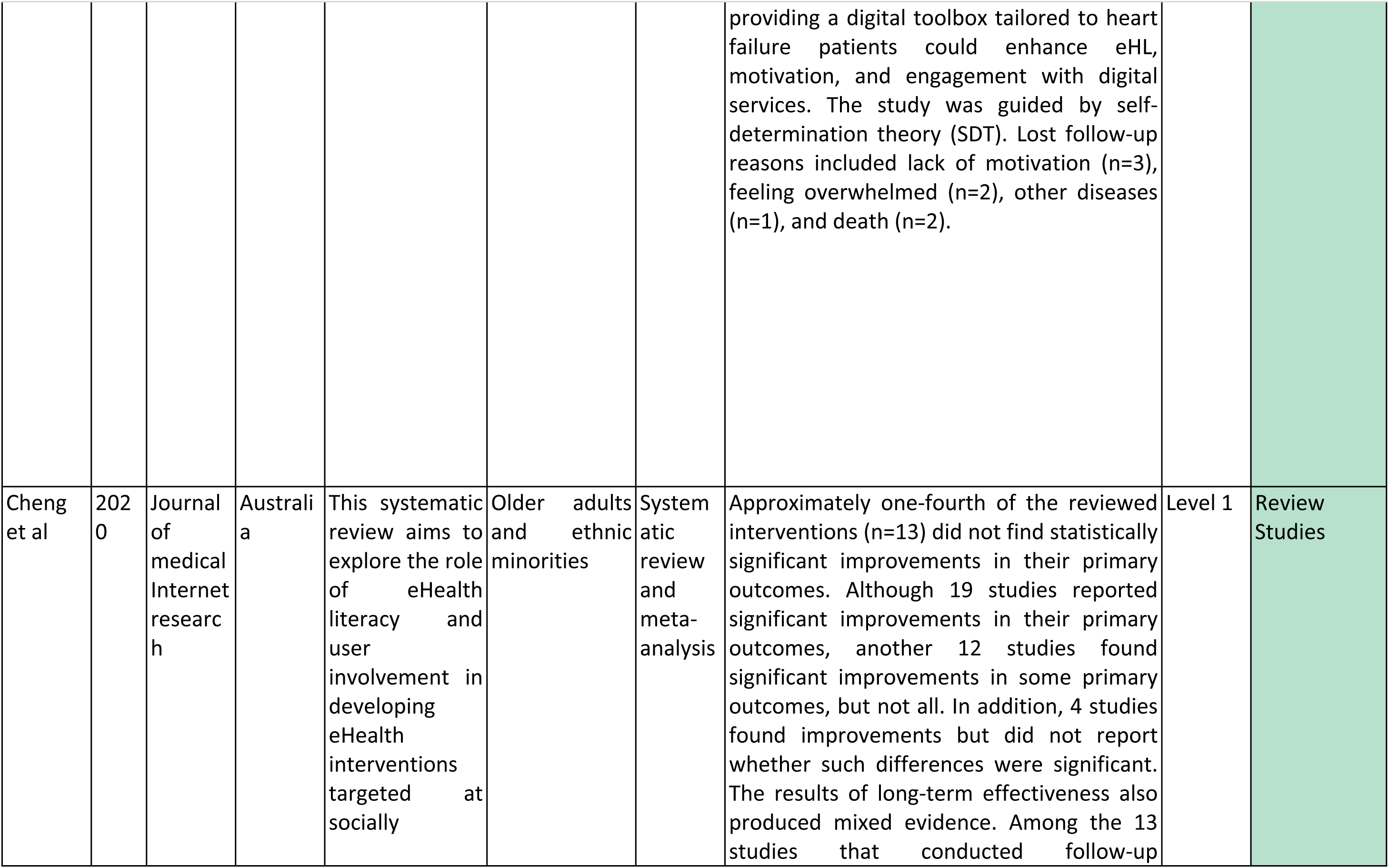
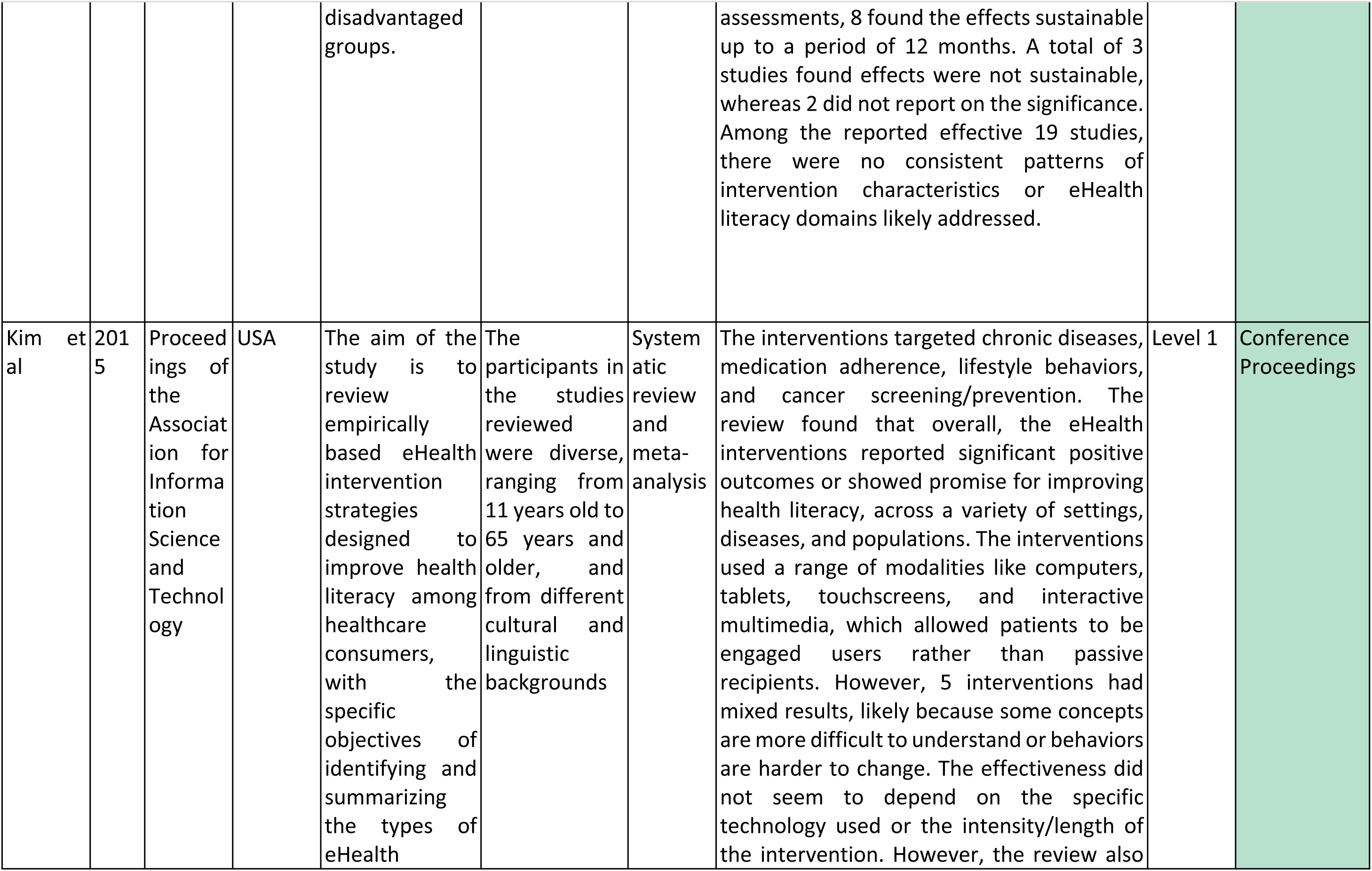
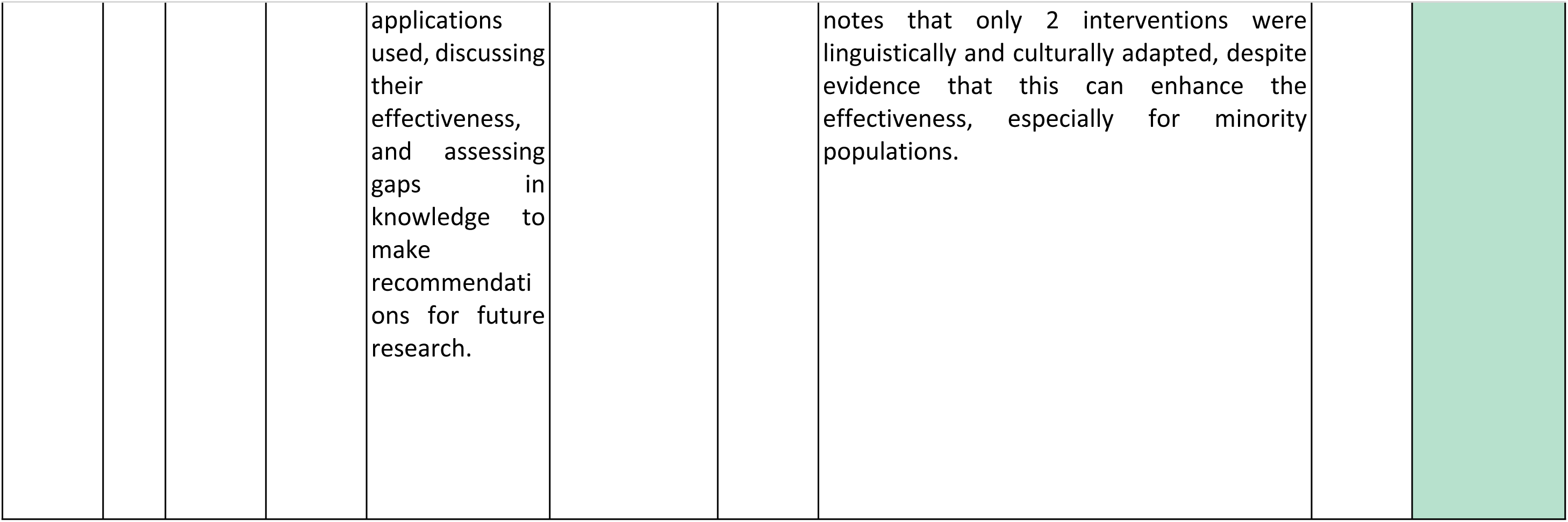
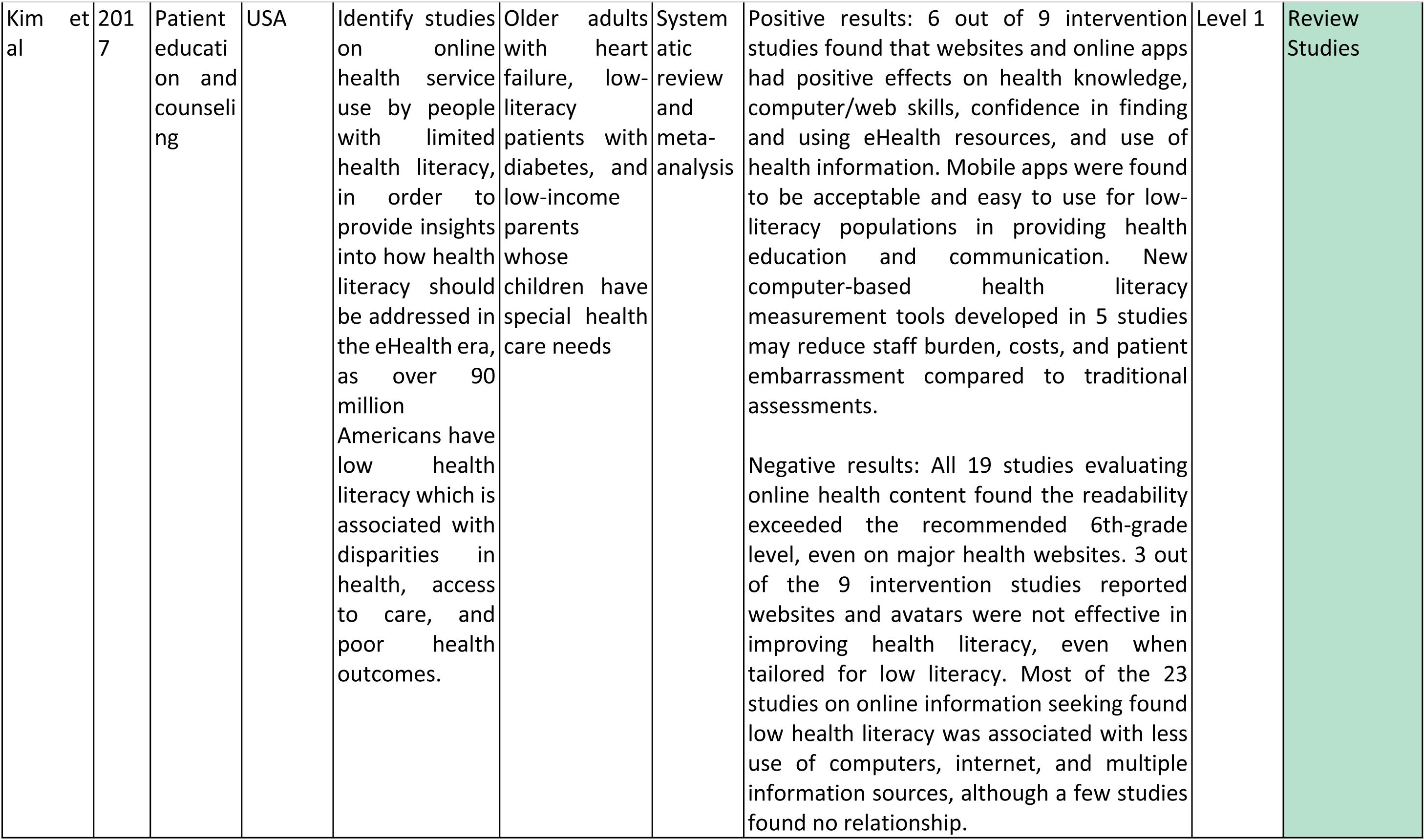
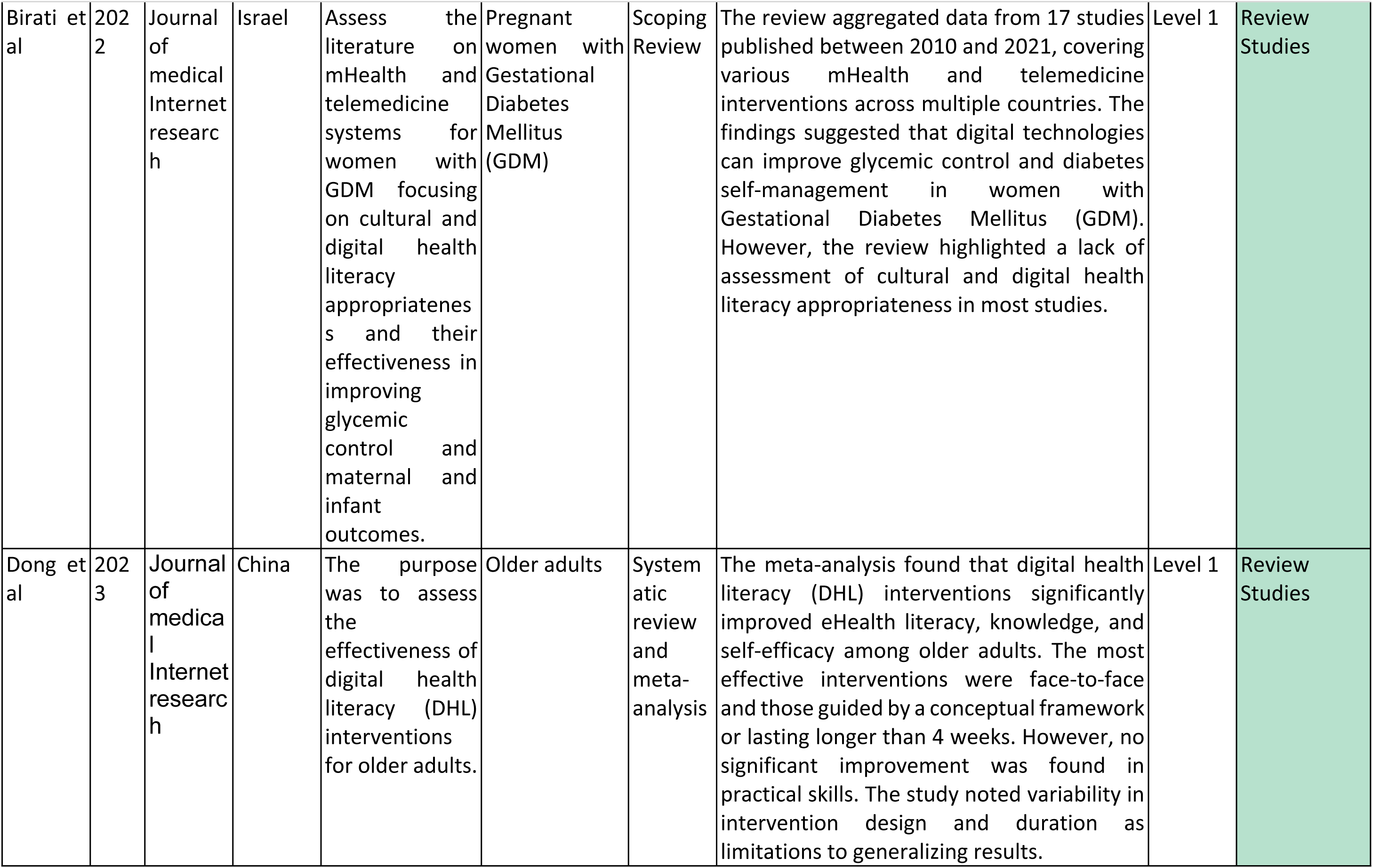
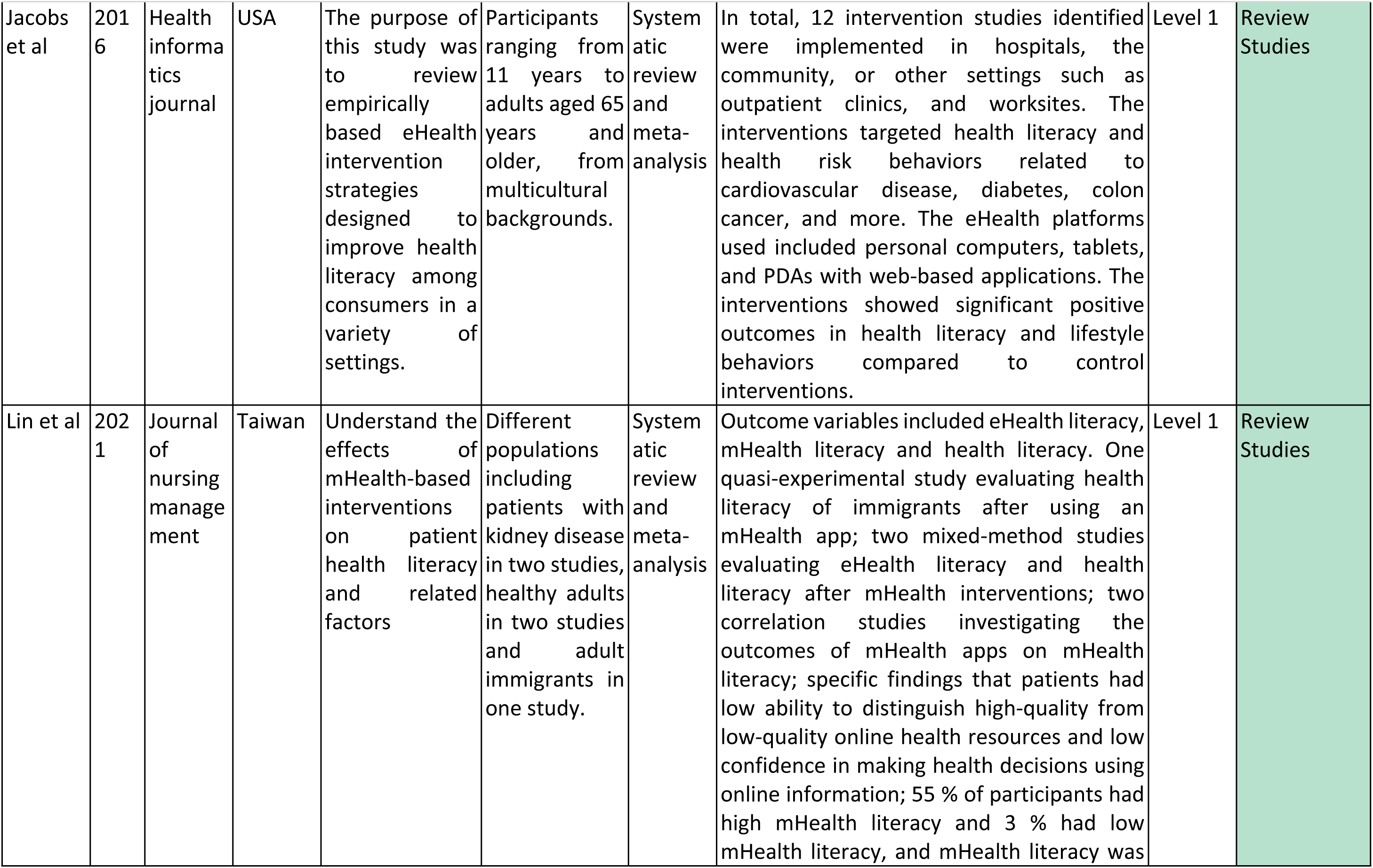
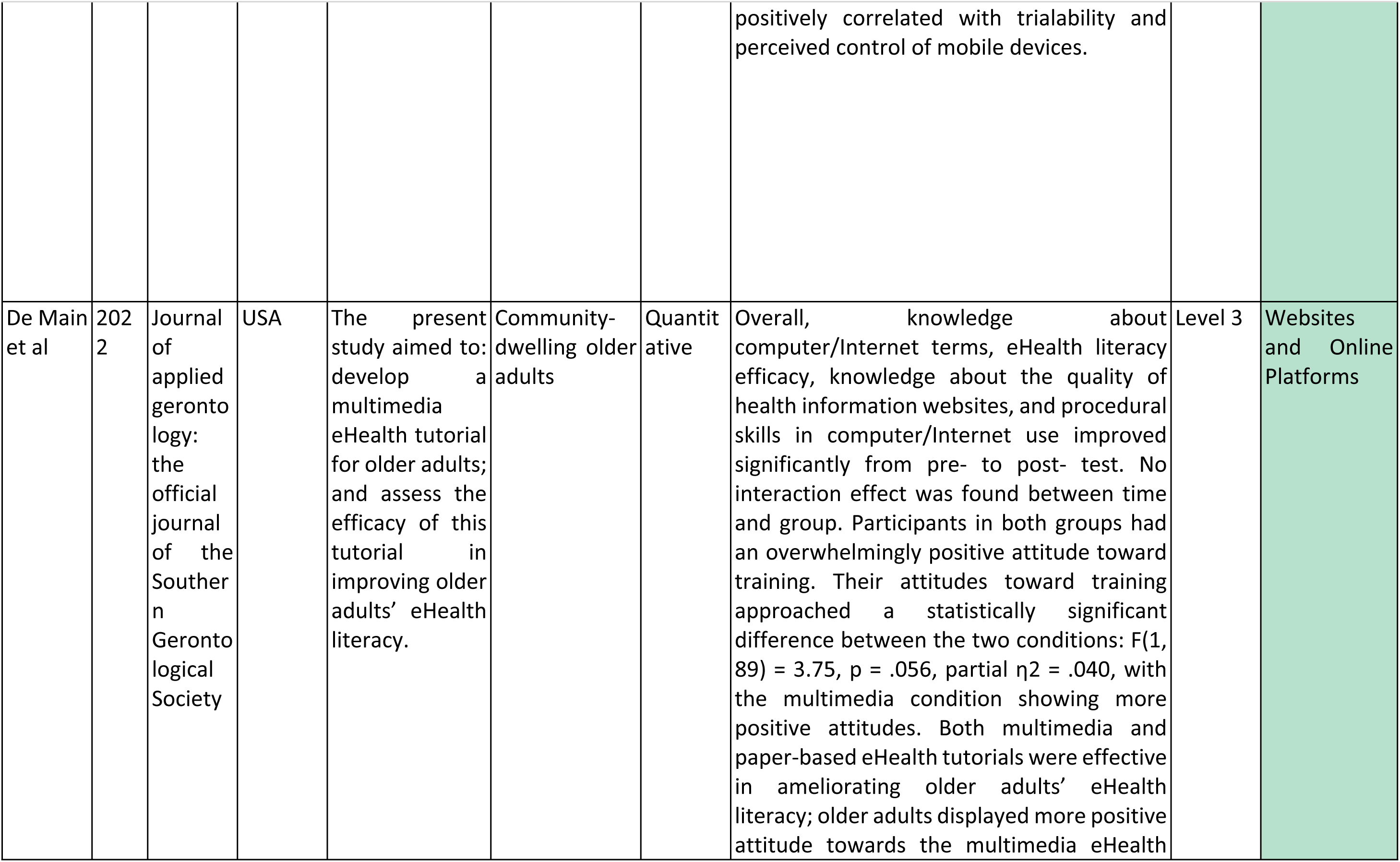
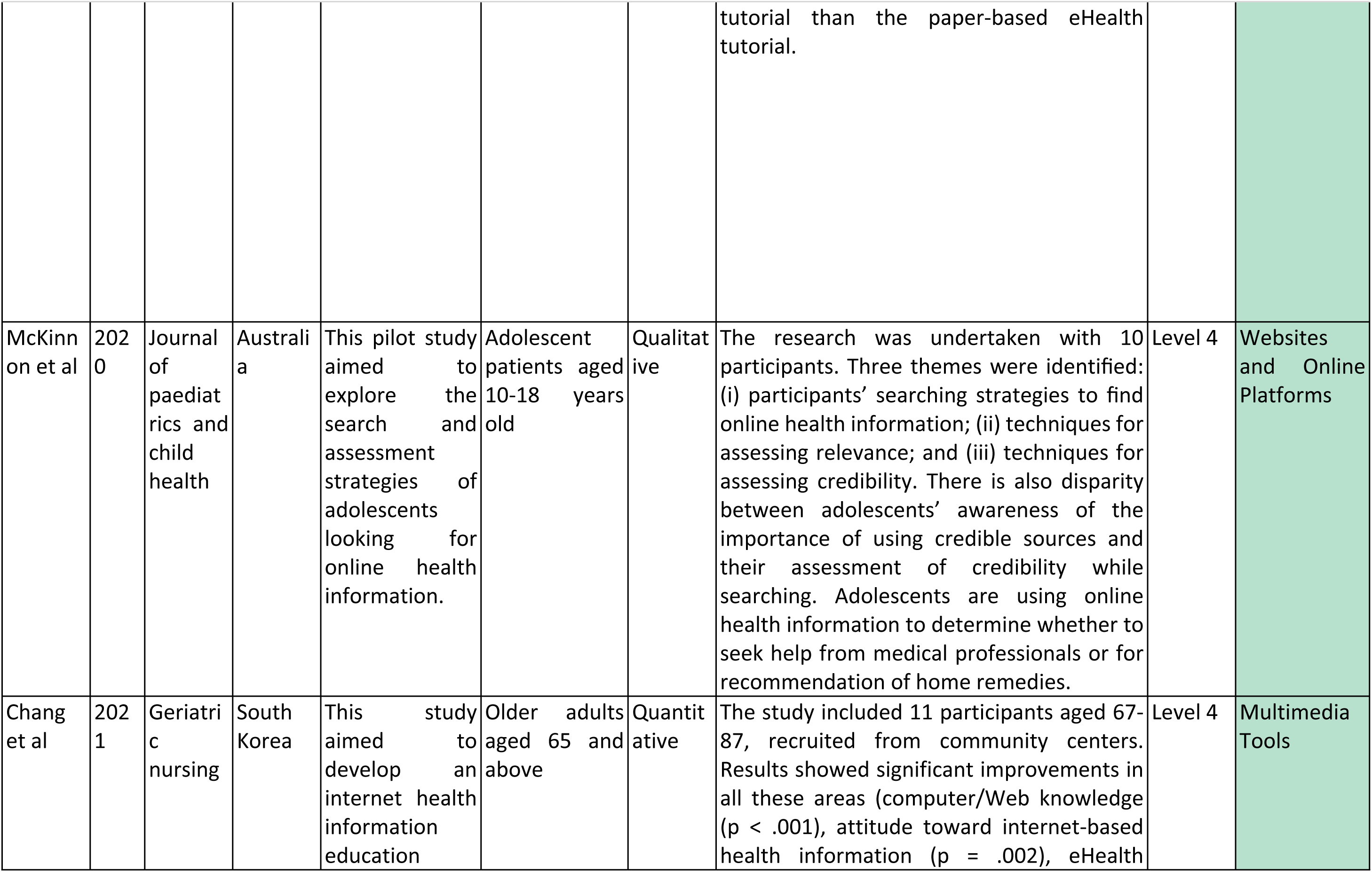
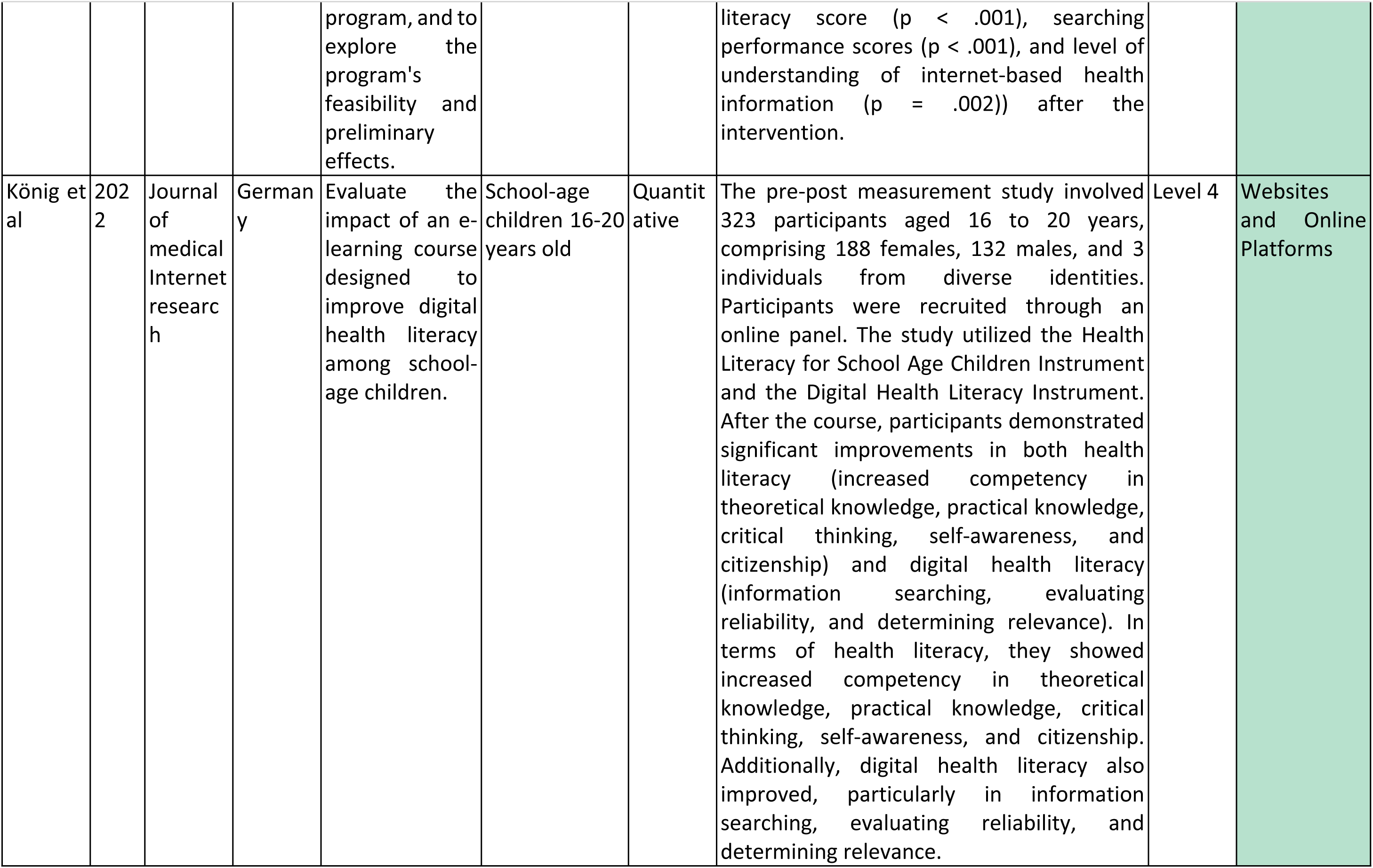
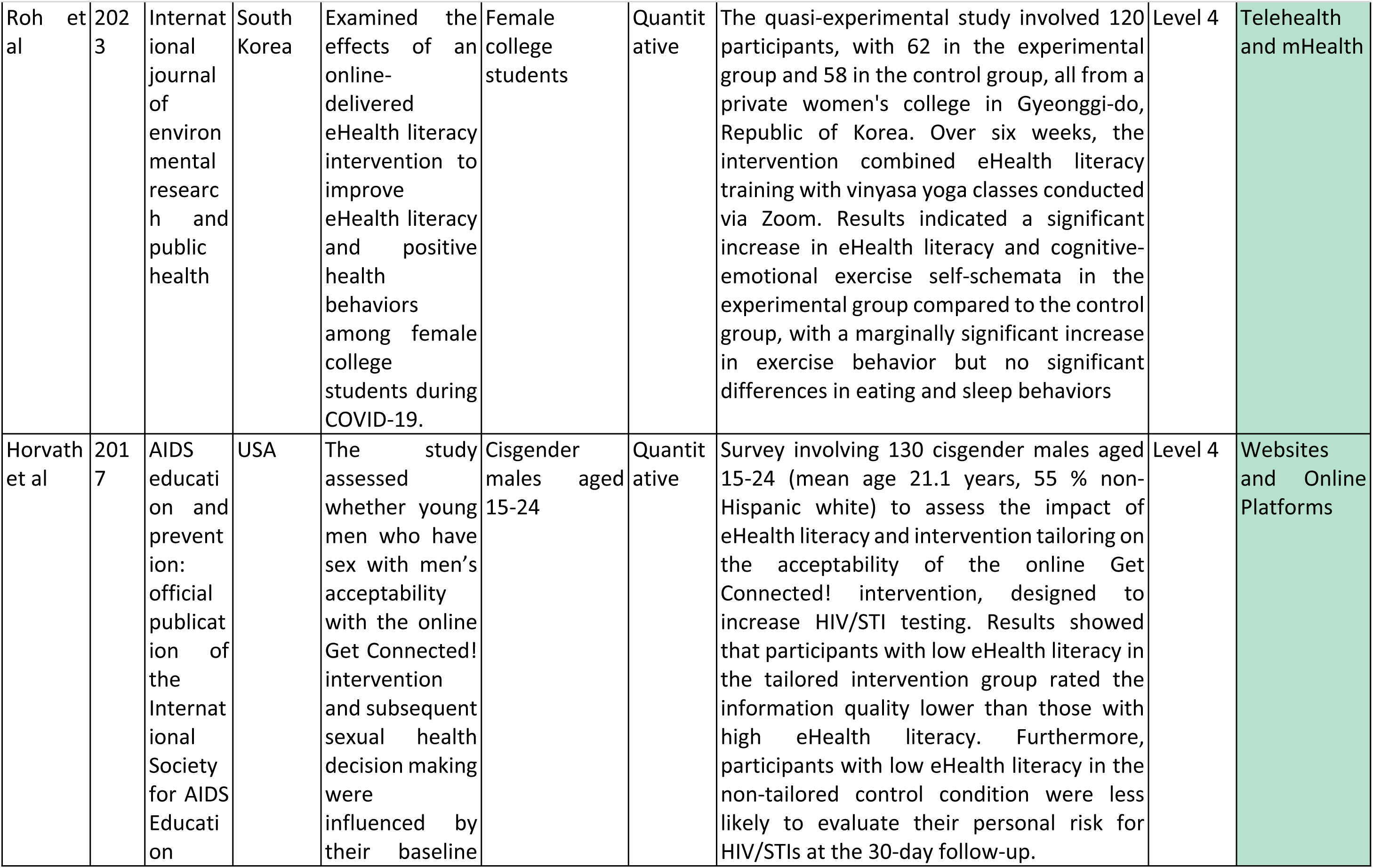
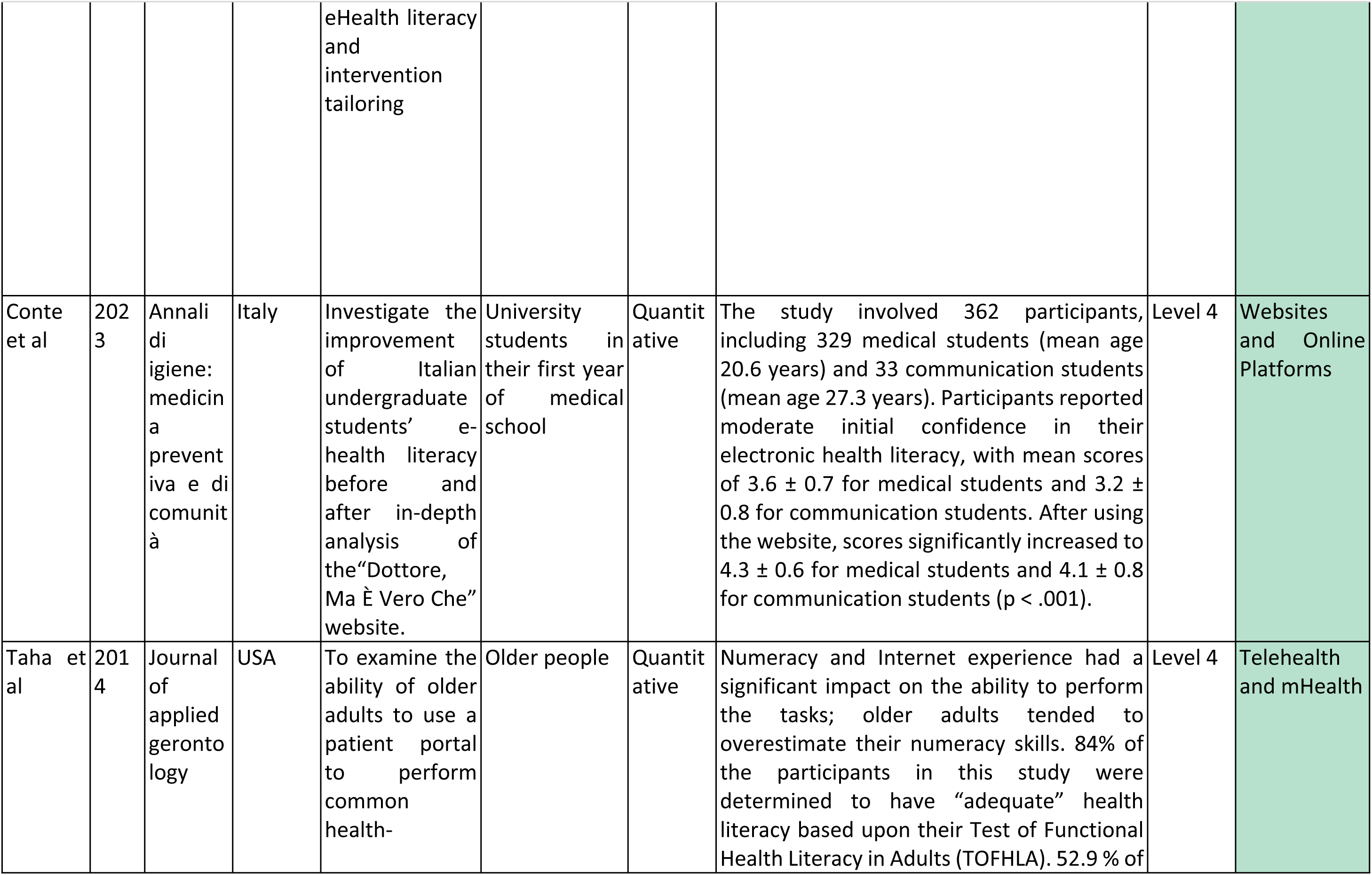
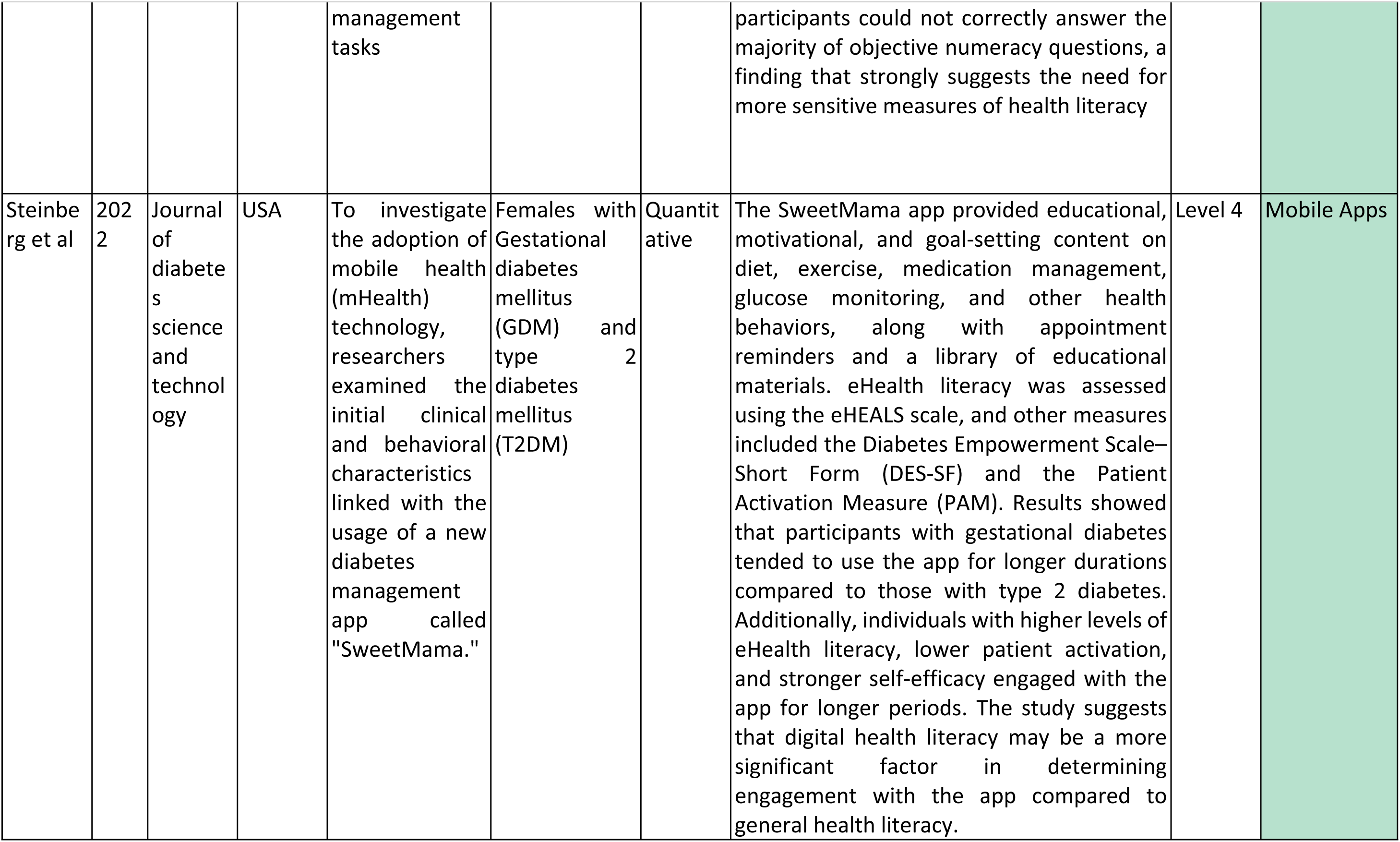
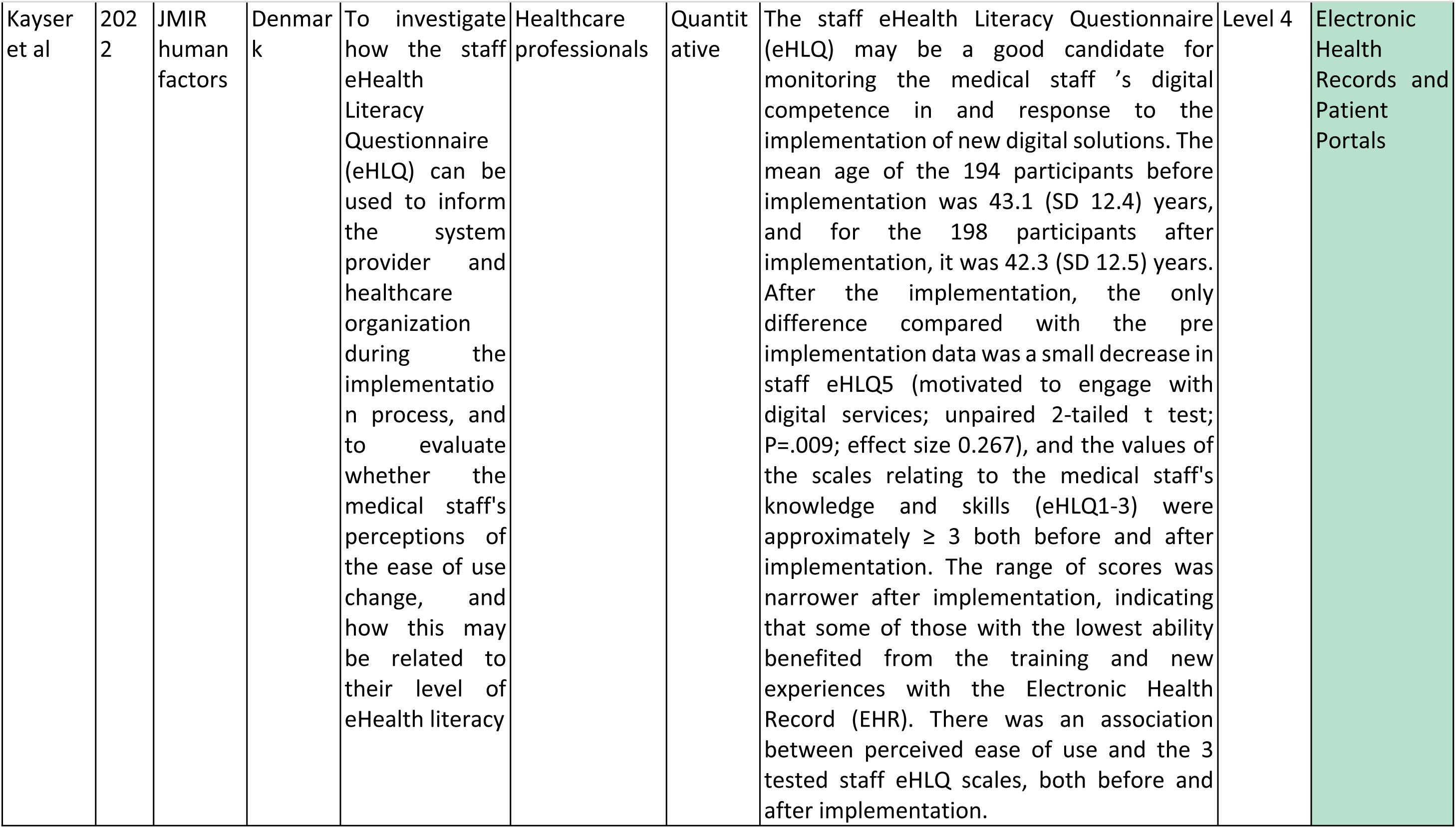
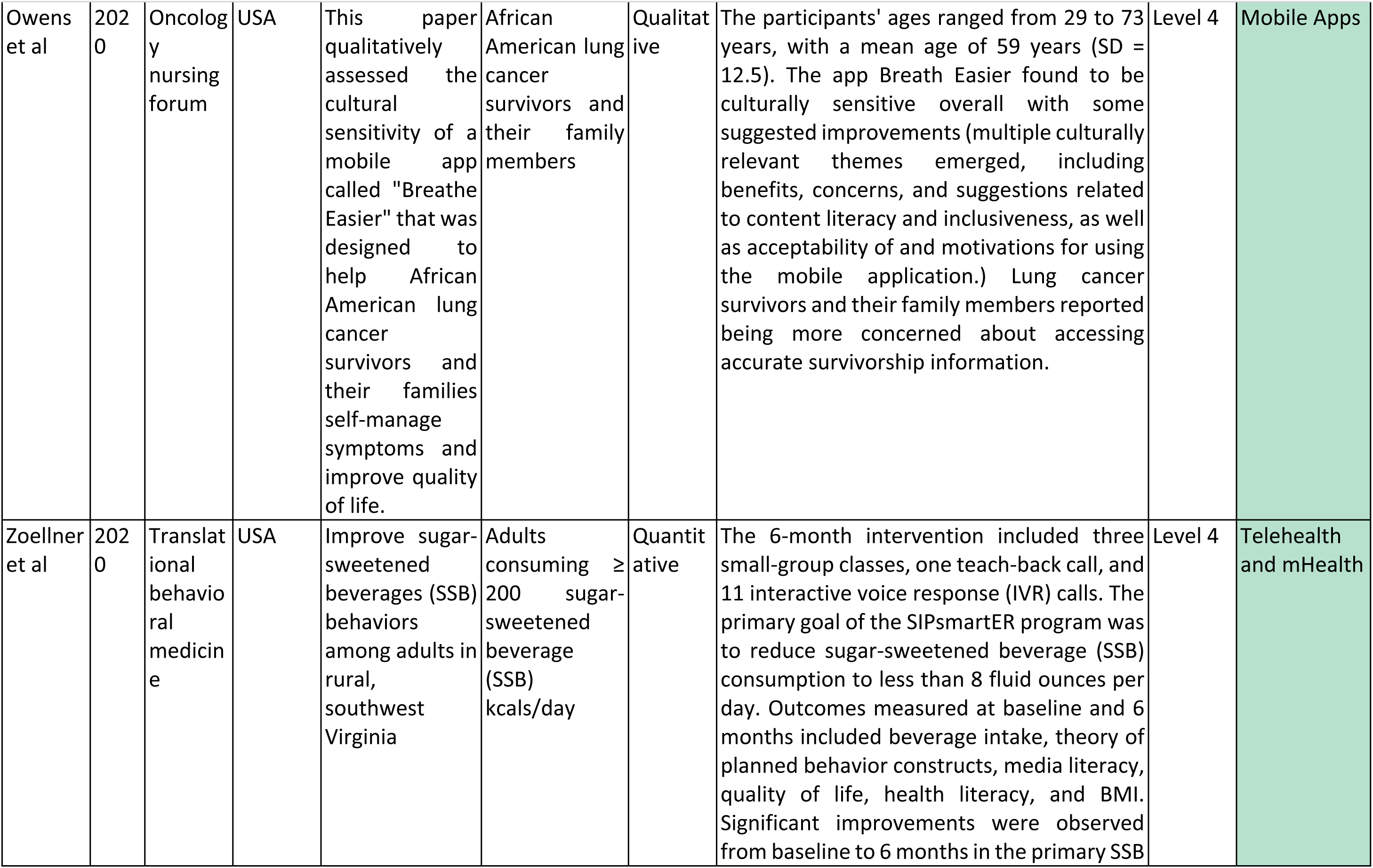
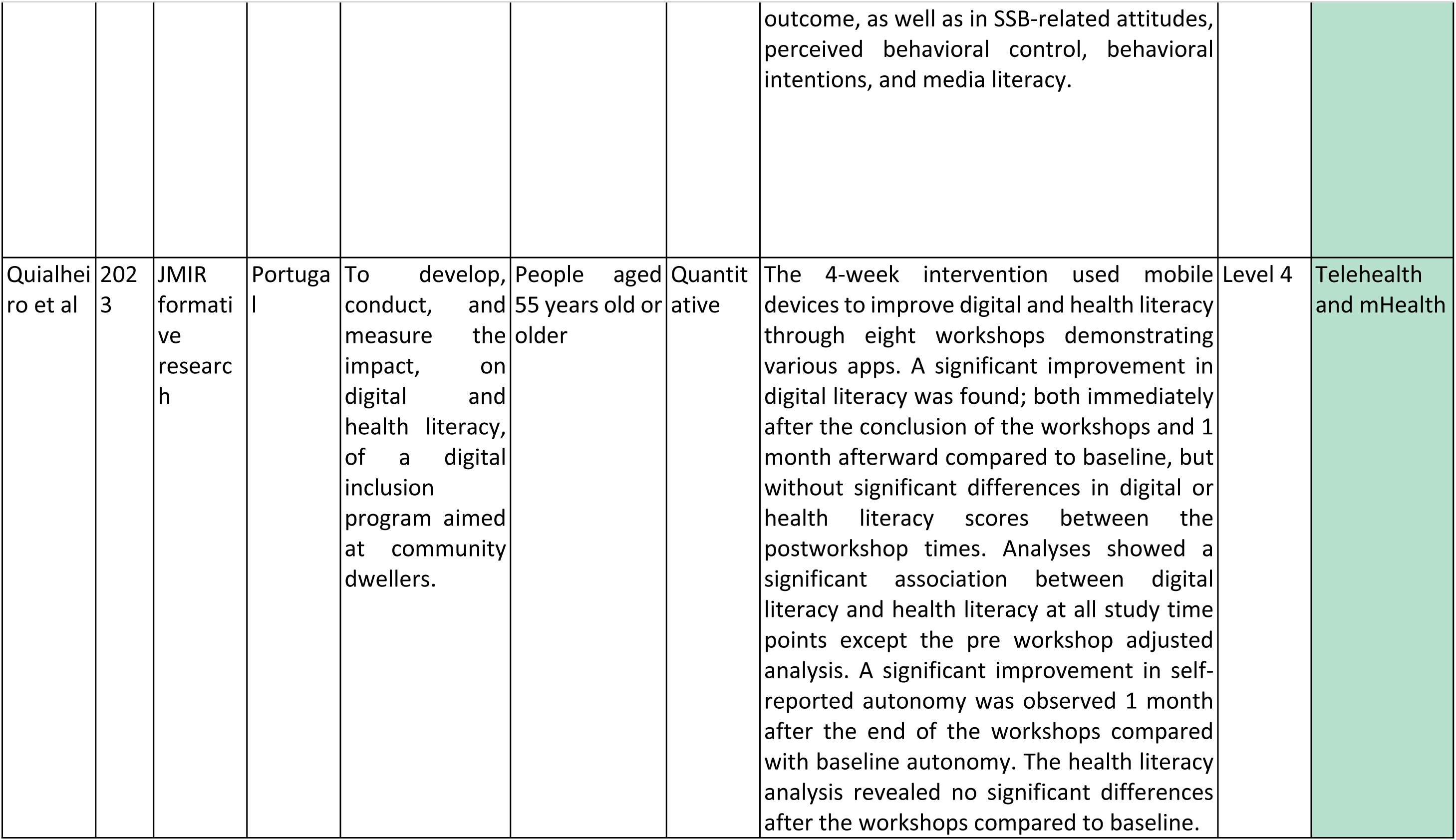
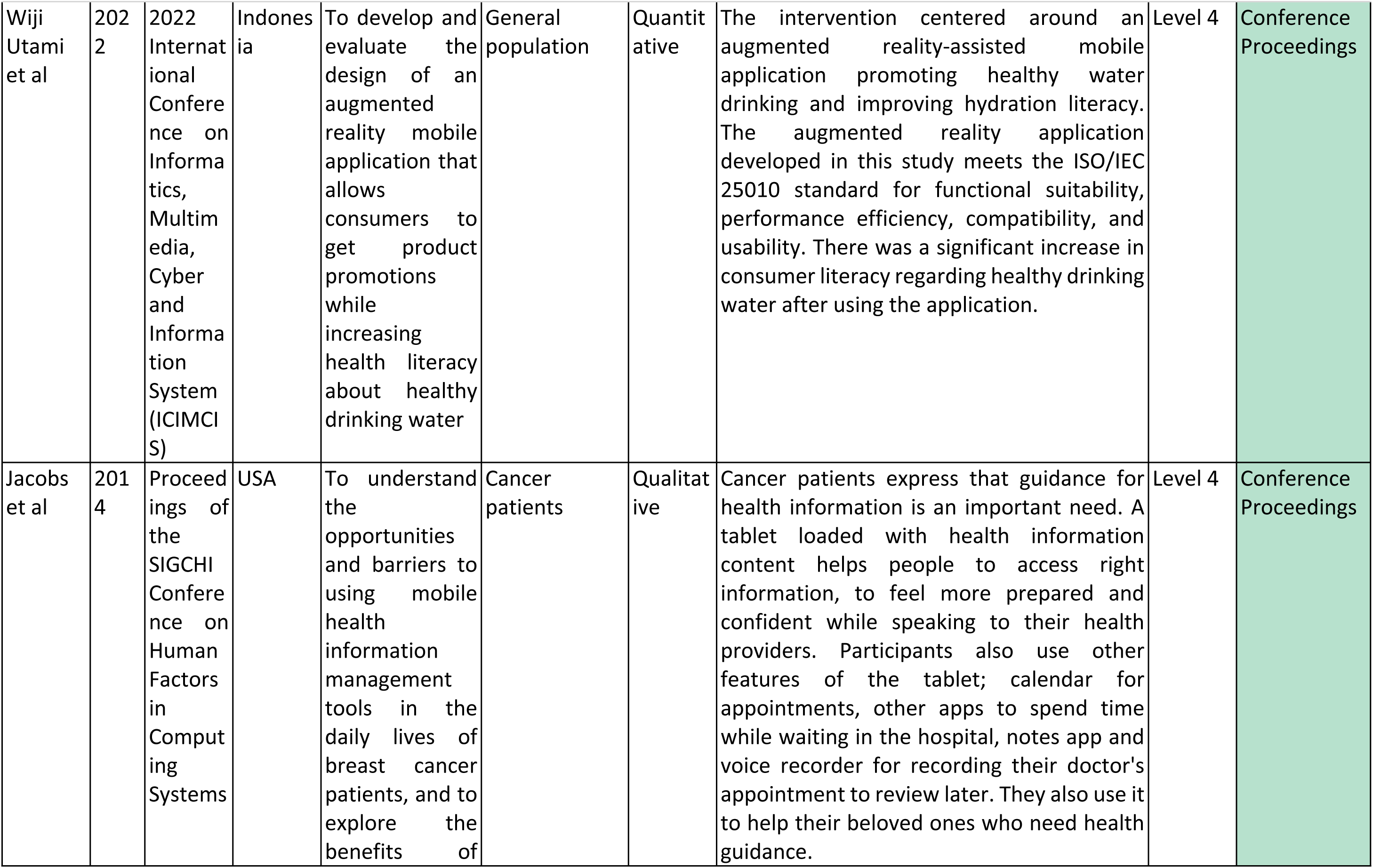
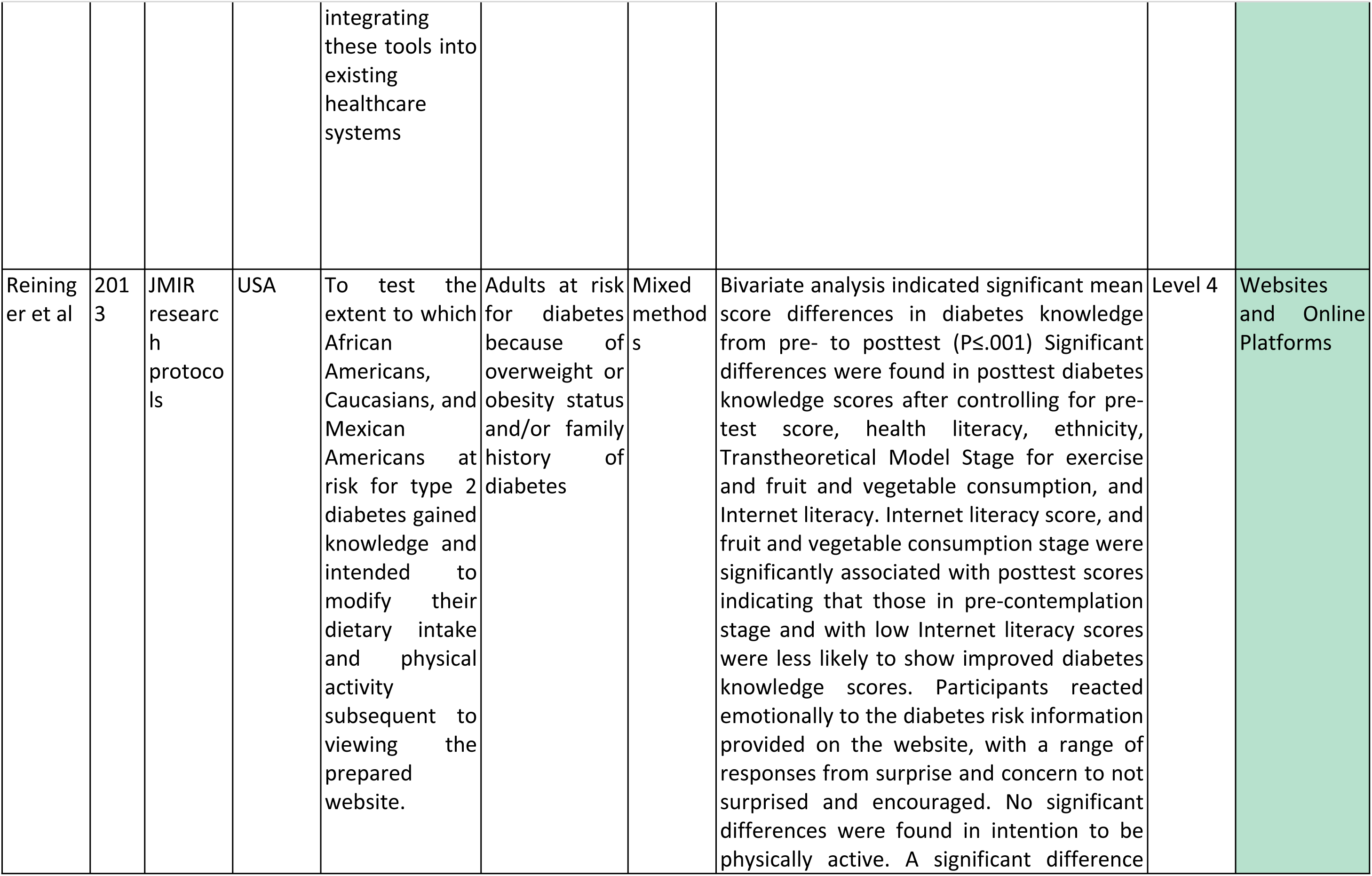
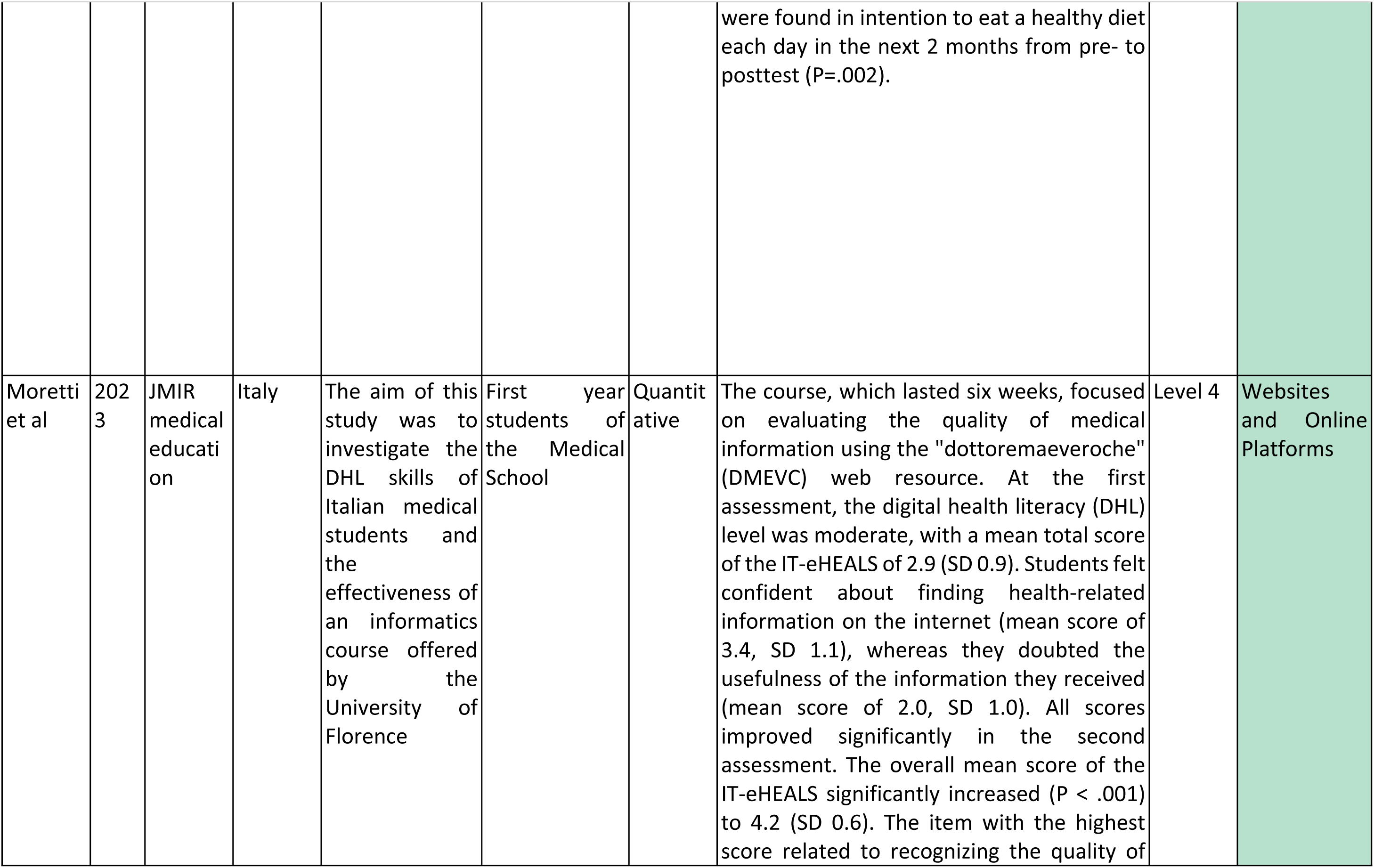
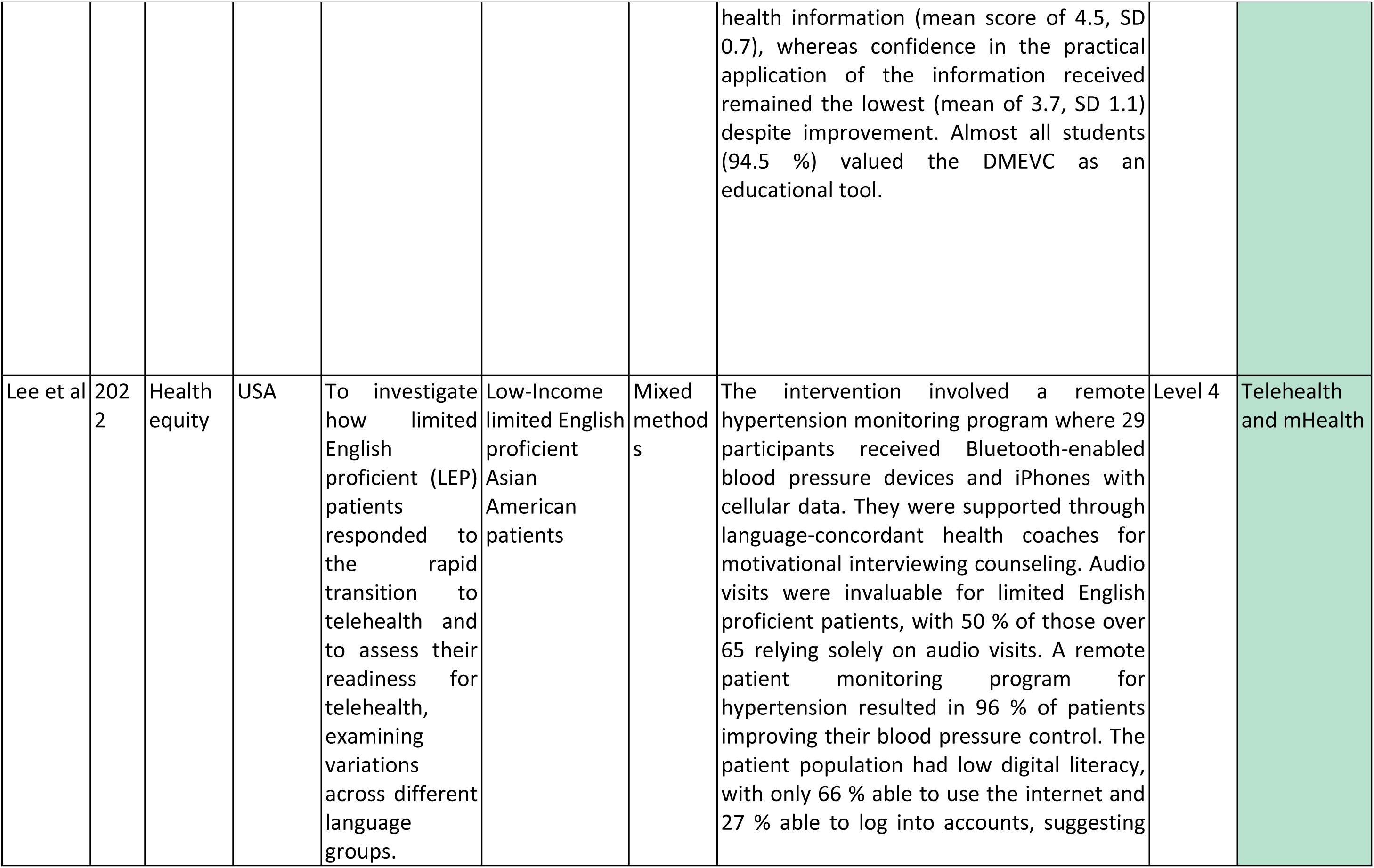
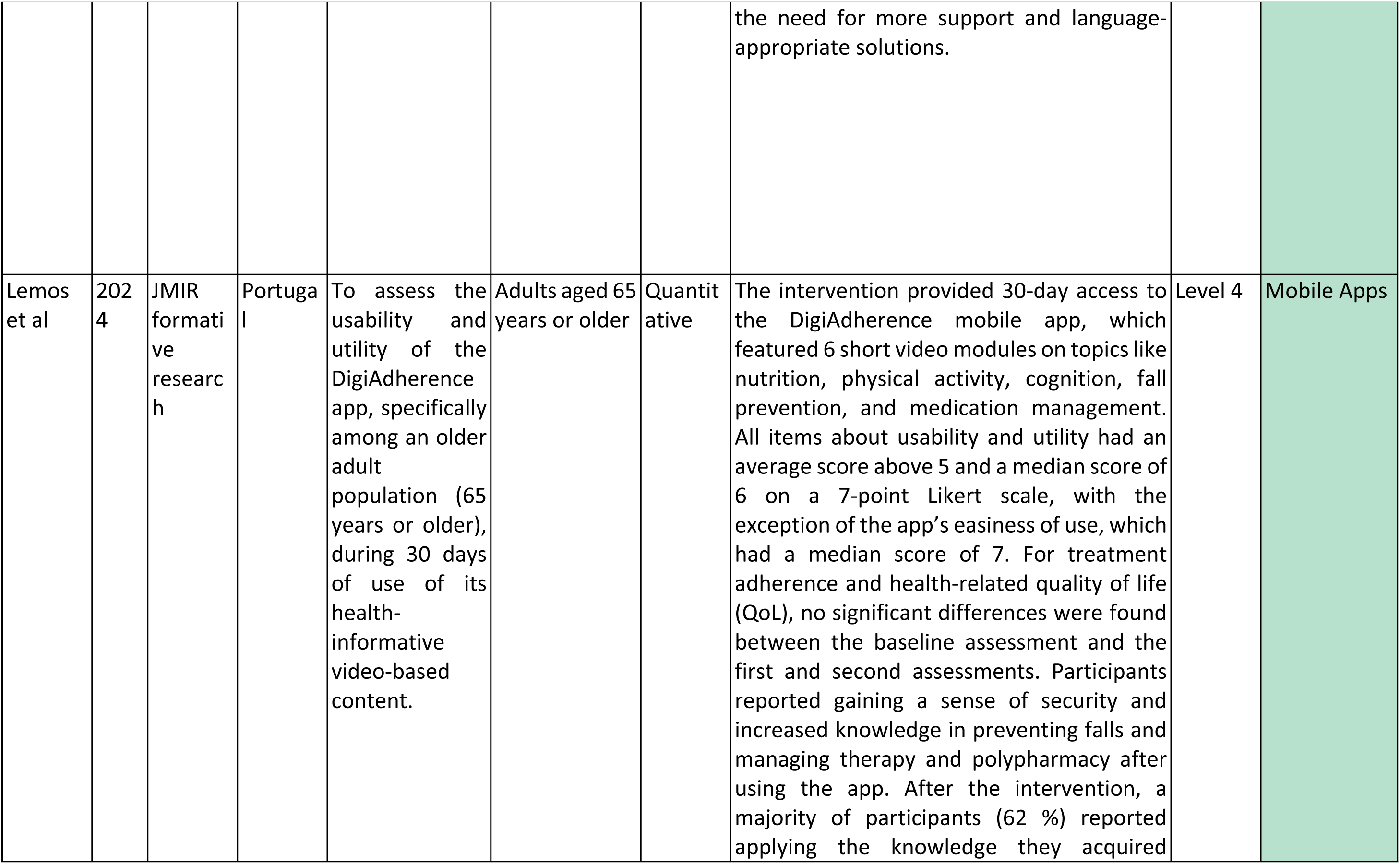
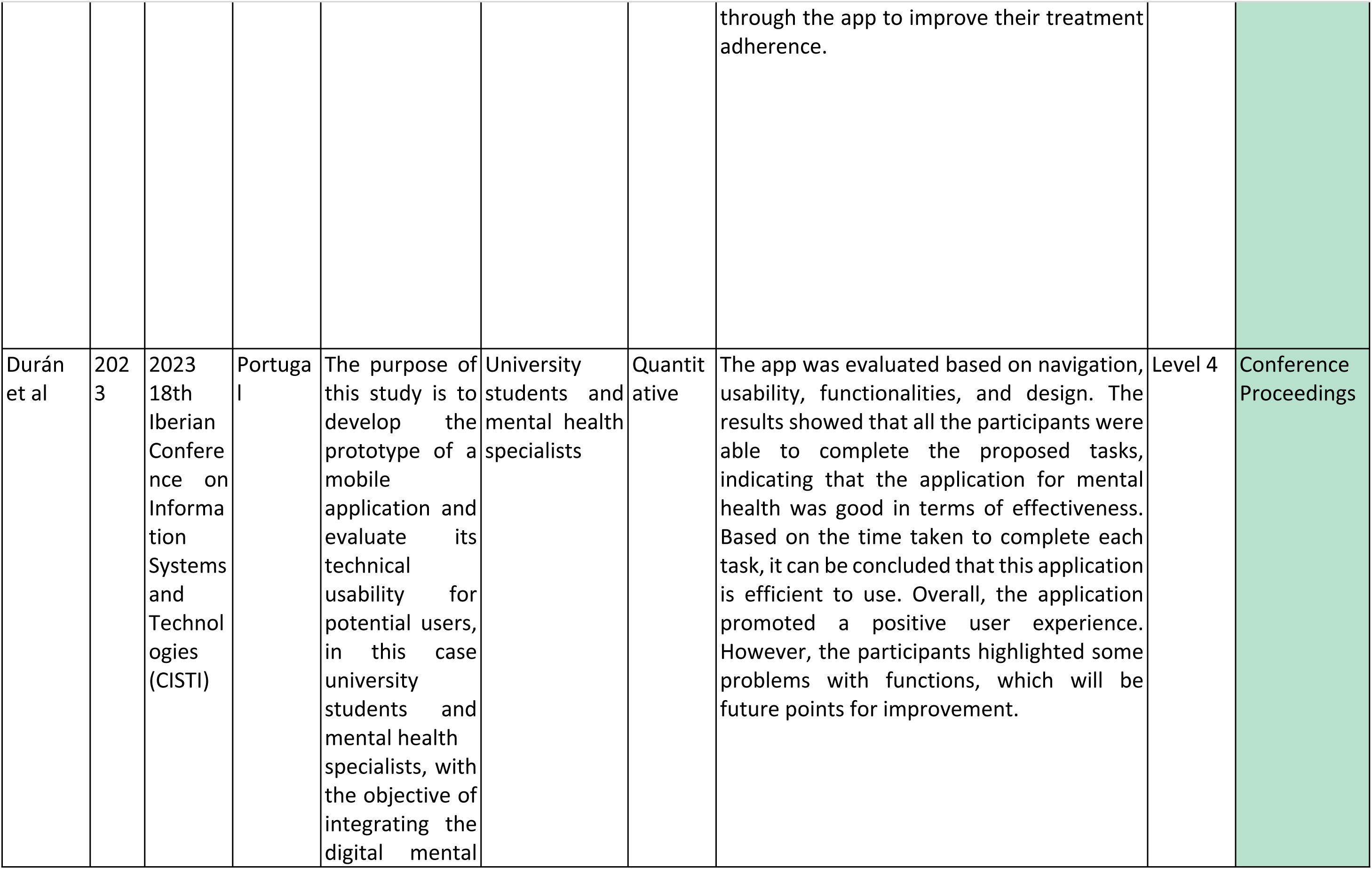
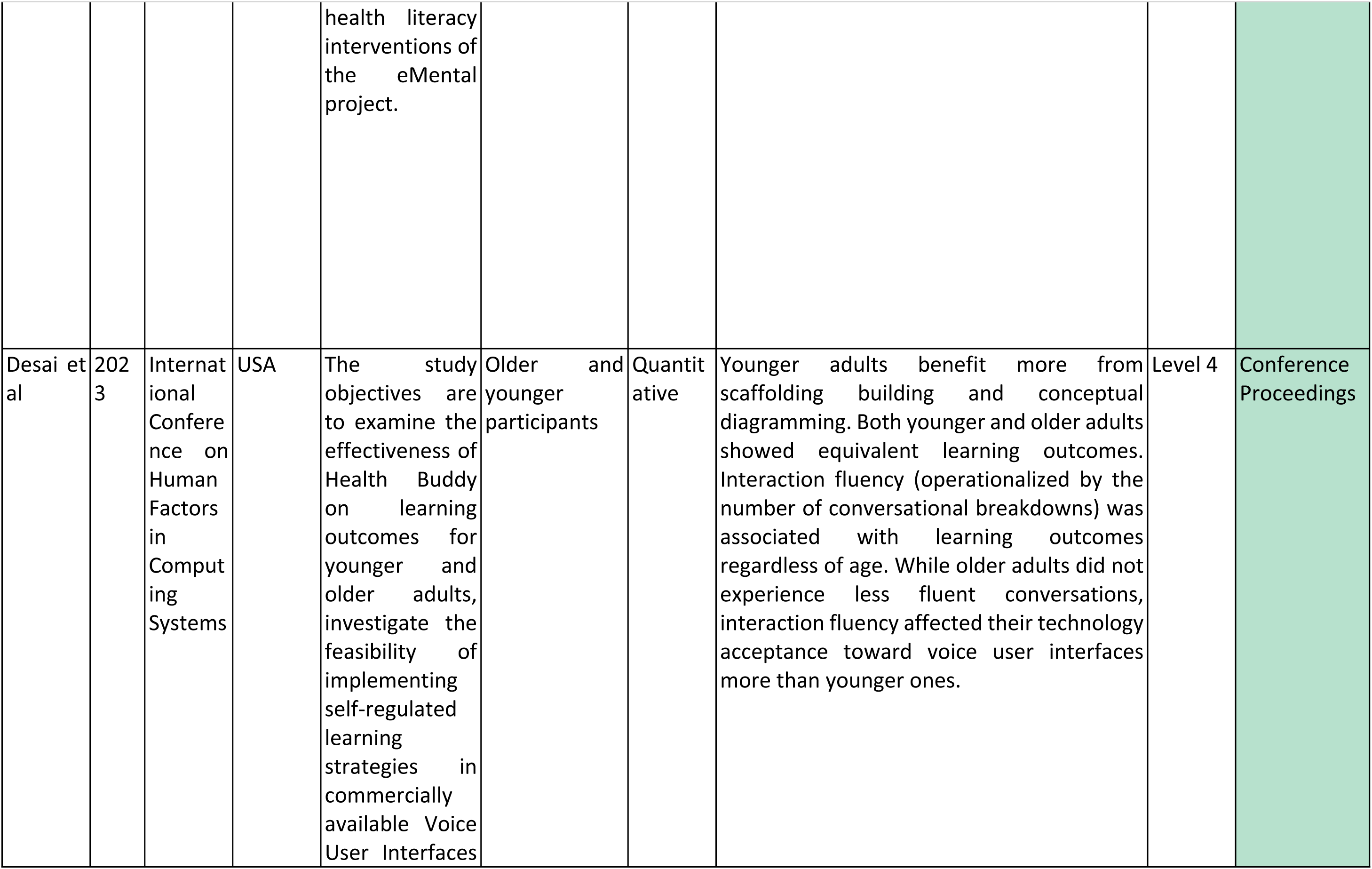
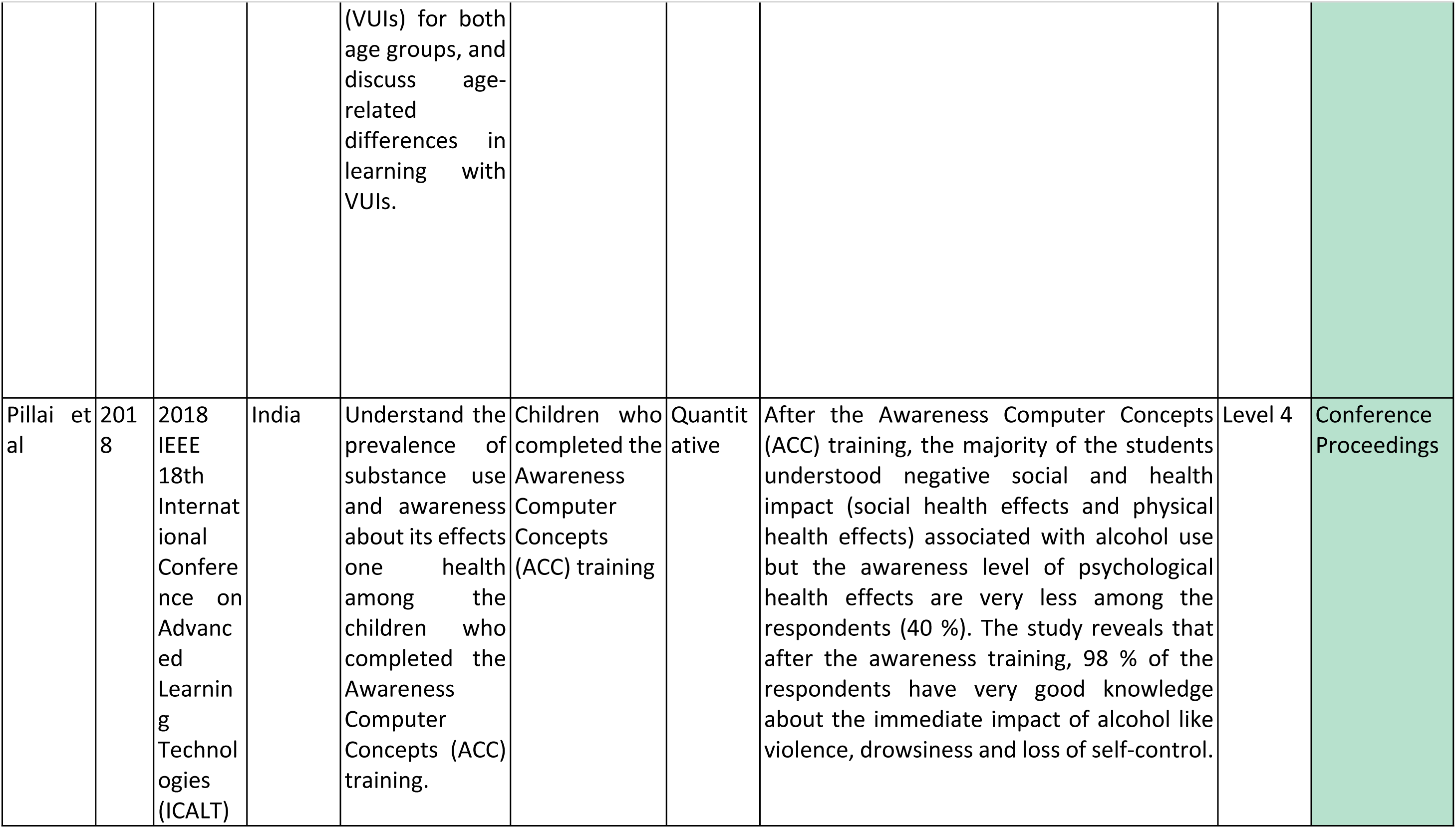
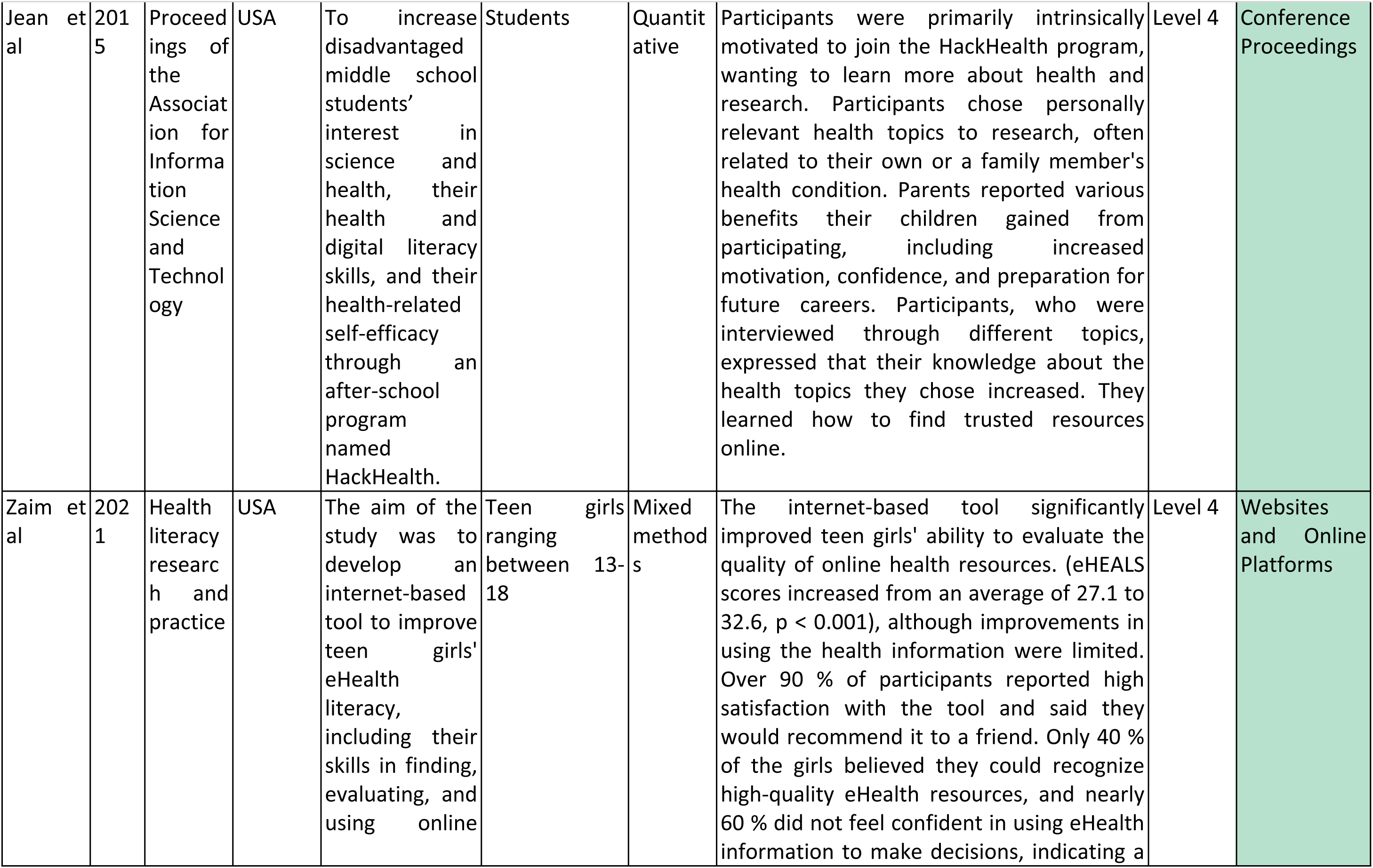
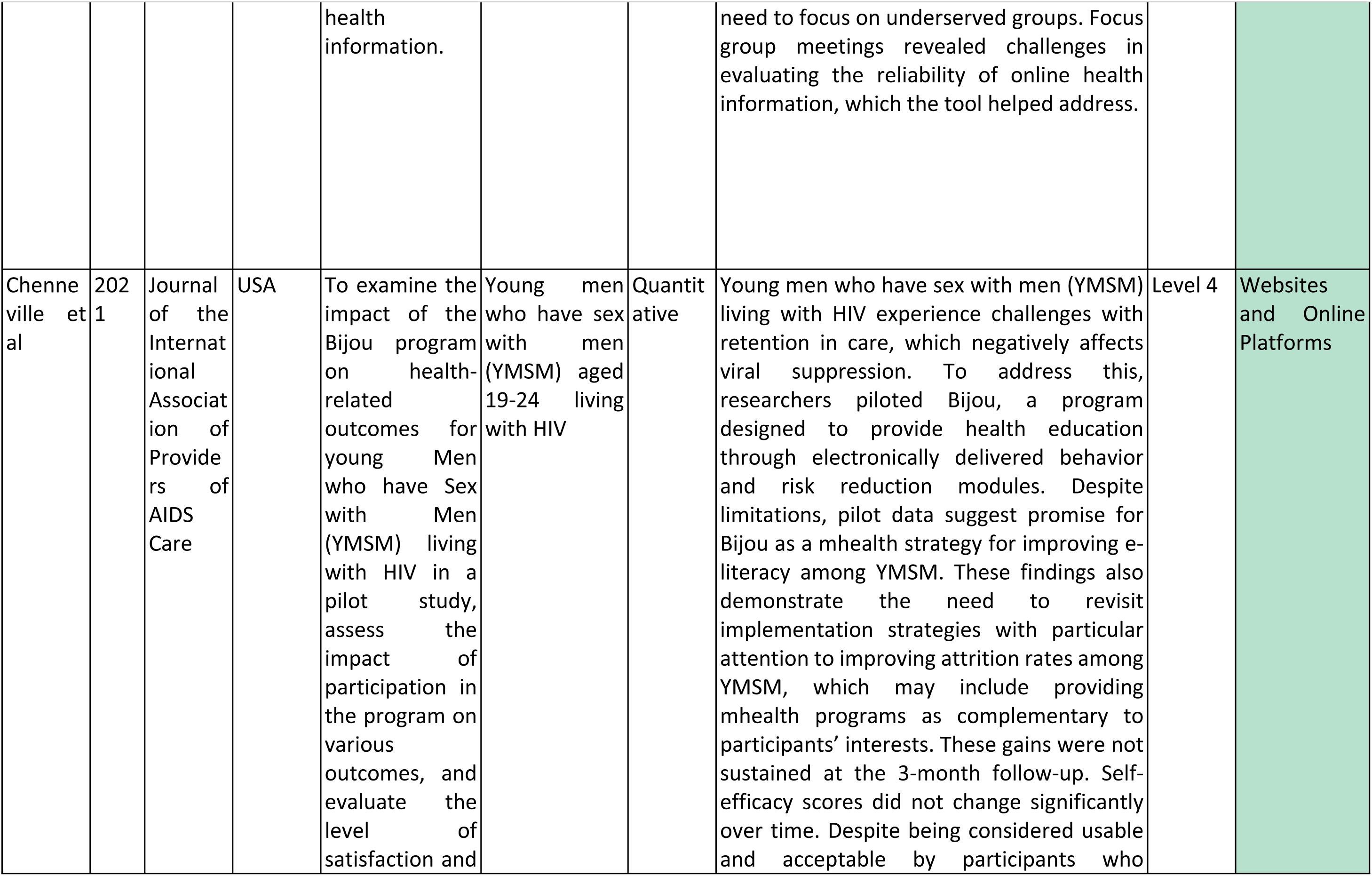
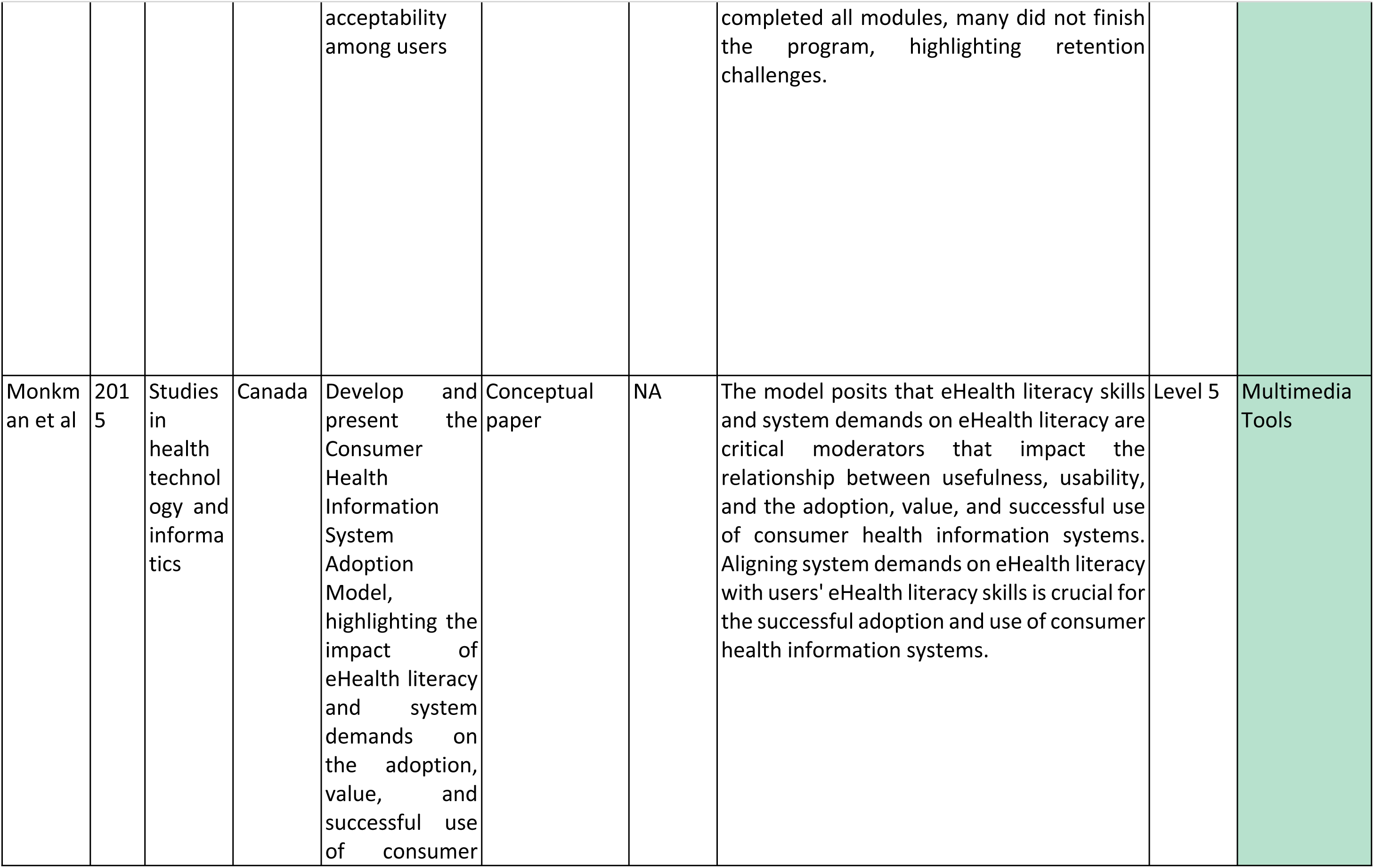
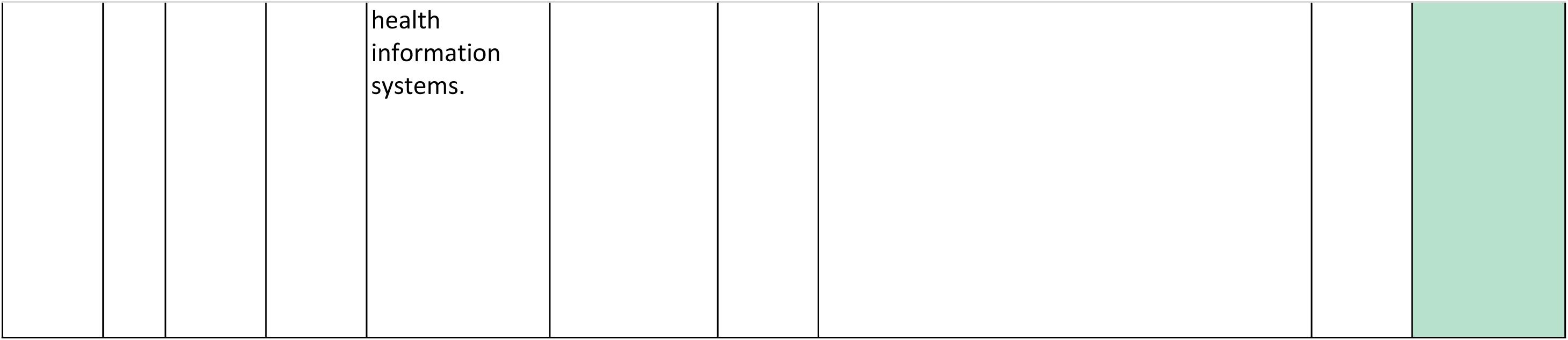
Overall characteristics of the studies included.

### 2.5. Reporting

We followed the Preferred Reporting Items for Systematic Reviews and Meta-Analyses (PRISMA) guidelines [22] to ensure comprehensive and transparent reporting of our review findings.

### 2.6. Data Analysis

We conducted a thematic analysis using data categorized from the Excel file. Where applicable, baseline characteristics were summarized as counts (N°) and percentages (%). Due to the heterogeneous nature of the retrieved studies, a meta-analysis was not conducted.

### 2.7. Ethical Considerations

This study did not require ethical approval as it involved reviewing published literature without direct interaction with human subjects.

## 3. Results

### 3.1. Main findings

While we identified 39 studies investigating various digital approaches to improve health literacy, the geographical distribution and population focus of these studies highlights significant disparities in research attention and implementation. The marked concentration of studies in North America (53.8 %, n = 21), particularly in the United States (n = 19), while understandable given the technological infrastructure and research funding available, raises concerns about the generalizability of findings to other contexts. The limited representation of studies from Europe (20.5 %, n = 8), Asia (17.9 %, n = 7), and Australia (7.7 %, n = 3), and notably, the complete absence of studies from Africa and South America suggests a significant gap in understanding how digital health literacy interventions perform in diverse healthcare systems and cultural contexts. This is further illustrated by tools such as the Global Digital Health Monitor, which shows that certain regions of the world lag in digital health adoption areas such as services and applications, strategy and investment, and workforce [23].

The age distribution of study populations also reveals important limitations. While adult populations were well-represented (33.3 %, n = 13), studies focusing specifically on elderly populations (10.3 %, n = 4) and young populations (12.8 %, n = 5) were relatively scarce. This is particularly concerning because these age groups often face unique challenges in digital literacy and healthcare access. The high proportion of studies with mixed or unspecified age groups (43.6 %, n = 17) further complicates our understanding of age-specific intervention effectiveness. The limited focus on ethnic minorities and underserved populations (17.9 %, n = 7) is another critical gap, especially considering that these groups often experience greater health disparities and may face additional barriers to digital health literacy [24]. The fact that most studies (69.2 %, n = 27) did not specify ethnic breakdown or included mixed populations without detailed analysis makes it difficult to understand the effectiveness of these interventions across different cultural and ethnic contexts. In terms of intervention types, 10.3 % of studies focused on mobile applications (n = 4) and 30.8% on websites and online platforms (n = 12), making these the most well-represented categories. In contrast, other potentially promising approaches received comparatively less attention, including multimedia tools (5.1 %, n = 2), telehealth and mHealth (15.4 %, n = 6), and electronic health records and patient portals (2.6 %, n = 1). Additionally, 17.9 % of studies were conference proceedings (n = 7), and another 17.9 % were review studies (n = 7).

Lastly, the majority of studies were classified as level 4 evidence (61.5 %, n = 24), indicating low-quality studies. Level 1 evidence accounted for 33.3 % (n = 13), while levels 3 and 5 were less common (2.6 % each, n = 1). Detailed study characteristics and evidence levels are presented in Table 4.

### 3.2. Narrative overview of the studies included

Next, we present a narrative overview of the included studies. The use of the narrative method offers a comprehensive overview of topics identified, facilitating a broader examination of subjects, while incorporating different perspectives and interpretations of the literature. This flexibility allows to summarize existing knowledge, identify gaps, and suggest future research avenues. By combining qualitative and quantitative studies, the narrative methodology can also uncover trends and patterns that might not be apparent in more structured review formats [25]. Therefore, interventional studies are divided according to the category of DHI/digital intervention used, while conference proceedings and reviews are presented in their own dedicated section as they generally discussed more thematic findings on DHI and health literacy. The final classification comprises seven categories:

1. **Mobile Applications**. Studies exploring health literacy interventions delivered via mobile apps (n = 4, 10.3 %) [26–29].
2. **Websites and Online Platforms**. Digital tools designed to disseminate health information or facilitate interactive learning experiences (n = 12, 30.8 %) [30–41].
3. **Multimedia Tools**. Interventions leveraging various media formats to enhance health literacy (n = 2, 5.1 %) [42,43].
4. **Telehealth and mHealth**. Remote healthcare services and mobile health technologies aimed at improving patient education and engagement (n = 6, 15.4 %) [44–49].
5. **Electronic Health Records (EHRs) and Patient Portals**. Studies assessing the role of digital health records in enhancing patient involvement and literacy (n = 1, 2.6 %) [50].
6. **Conference Proceedings**. Research presented at conferences, offering valuable insights into emerging trends and innovations (n = 7, 17.9 %) [51–57].
7. **Review Studies**. Systematic and narrative reviews synthesizing existing evidence in the field (n = 7, 17.9 %) [12,13,15,58–61].

#### Mobile Apps

Mobile app interventions represent a significant portion of applications addressing health literacy to improve chronic conditions, with 4 studies focusing on various populations and health conditions. *Owens et al. (2020)* [26] implemented a mobile app designed for symptom self-management and quality of life improvement in African American lung cancer survivors and their families. The app was perceived as culturally sensitive and inclusive. *Parker et al. (2022)* [27] targeted overweight and obese adults with a mobile app-based health coaching, showing improvements in diet-related health literacy but limited effects on physical activity. *Steinberg et al. (2022)* [28] evaluated the SweetMama app for diabetes in pregnant women, finding varying levels of engagement based on eHealth literacy. *Lemos et al. (2024)* [29] demonstrated high user satisfaction with a mobile app for older adults, though device usage skills were limiting.

#### Websites and Online Platforms

This category represents the largest group with 12 studies examining various web-based interventions. *Chenneville et al.* (2021) [30] examined the challenges faced by young men who have sex with men (YMSM) living with HIV, particularly in maintaining engagement with care, a crucial factor for achieving viral suppression. The Bijou mHealth program showed promise in improving e-literacy but struggled with high dropout rates, limiting its long-term impact. To enhance retention and ensure sustainable benefits, more effective strategies are needed. Similarly, *Reininger et al.* (2013) [31] explored a diabetes prevention website across different ethnic groups, finding that while participants gained knowledge, internet literacy varied significantly among them. *Horvath et al.* (2017) [32] investigated the impact of tailored online interventions on LGBTQ youth, revealing positive effects on their sexual health decision-making.

*McKinnon et al.* (2020) [33] studied how adolescents searched for health information online, demonstrating that while they could easily access information, they often failed to assess its credibility. Addressing this gap, *Zaim et al.* (2021) [34] developed the eTeen Health tool, which significantly improved teenage girls’ ability to evaluate online health resources. Likewise, *Scull et al.* (2021, 2022) [35,36] assessed a web-based program aimed at enhancing sexual health literacy among adolescents, reporting notable improvements in both media literacy and sexual health knowledge. *König et al.* (2022) [38] also contributed to this field, demonstrating how an e-learning course significantly boosted adolescents’ digital health literacy, as well as their subjective and objective health knowledge.

Focusing on older populations, *De Main et al.* (2022) [37] compared multimedia eHealth tutorials with traditional paper-based tutorials, finding that digital approaches led to better eHealth literacy. *Conte et al.* (2023) [39] extended this research to Italian university students, showing substantial improvements in digital health literacy following exposure to an educational website.

In a different approach, *Bickmore et al.* (2016) [40] evaluated a conversational search engine designed to assist individuals with low health and computer literacy in finding clinical trials. Their randomized trial compared this system to a conventional keyword-based search engine, concluding that conversational agents could improve accessibility and reduce recruitment bias against disadvantaged populations. Finally, *Moretti et al.* (2023) [41] demonstrated that Italian medical students enhanced their digital health literacy through an informatics course utilizing the “dottoremaeveroche” tool, emphasizing the importance of such resources for advancing public health communication and access to reliable health information.

#### Multimedia Tools

2 studies explored various multimedia approaches to health literacy. *Monkman et al. (2015)* [42] demonstrated that a conversational search engine improved satisfaction for cancer patients with low health literacy. *Chang et al. (2021*) [43] showed that internet health education programs improved digital literacy among seniors in South Korea.

#### Telehealth and mHealth

6 studies focused on telehealth and mobile health interventions. *Taha et al. (2014)* [44] found that although many older adults had adequate general health literacy, they struggled with the digital skills required for patient portal use. *Zoellner et al. (2020)* [45] used an interactive voice response system to reduce sugar-sweetened beverage intake, showing significant behavior change over six months. *Spindler et al. (2022)* [46] used telerehabilitation to enhance digital health literacy and engagement among heart failure patients, finding that heart failure patients retained high engagement with digital health tools over time. *Roh et al. (2023)* [47] combined eHealth literacy training with physical activity interventions, leading to improvements in both literacy and behavior. *Lee et al. (2022)* [48] explored telehealth readiness among limited English proficient Asian American patients, resulting in improved blood pressure control. *Quialheiro et al.* (2023) [49] developed and evaluated a digital inclusion program for community members, assessing its impact on digital and health literacy; participants improved digital literacy after 8 workshops and maintained their skills 1 month later, but health literacy showed no significant change during the project.

#### Electronic Health Records and Patient Portals

*Kayser et al. (2022)* [50] showed that healthcare professionals’ eHealth literacy remained stable after implementing an electronic health record system, emphasizing the need for continuous training.

#### Conference Proceedings

The conference proceedings analyzed in this section, predominantly from technology-focused venues such as IEEE and ACM symposiums, emphasize technical experimentation and design considerations of digital health interventions (DHIs) rather than longitudinal clinical outcomes. These studies often present preliminary implementations or proof-of-concept validations, offering insights into technological feasibility and user interface considerations that complement broader health literacy research. 7 conference proceedings were included in our search, although not followed by an original article.

*Jacobs et al. (2014)* [51] conducted a preliminary investigation of a mobile tool for cancer patients, who expressed a significant need for guidance on health information. *Jean et al. (2015)* [52] reported on the impacts of an after-school program aimed to increase disadvantaged middle school students’ health and digital literacy skills. At the end of the study, parents reported various benefits, including increased motivation, confidence, and preparation for future careers. *Desai et al (2023*) [53] examined the effectiveness of a Voice User Interface (VUI) for informal self-regulated learning of health topics among younger and older adults, where younger adults benefited more from scaffolding building and conceptual diagramming, while both age groups demonstrated equivalent learning outcomes. *Pillai et al (2018)* [54] after the Awareness Computer Concepts initiative, most students understood the social and physical health effects of alcohol, but awareness of psychological effects remained low (40%). However, 98% gained strong knowledge about alcohol’s immediate impacts, such as violence, drowsiness, and loss of self-control, while *Wiji Utami et al. (2022)* [55] focused on using augmented reality to promote hydration literacy. *Kim et al (2015)* [56] reviewed eHealth intervention strategies aimed at improving health literacy among healthcare consumers, suggesting that effective eHealth interventions for health literacy do not need to be extensive or expensive, and need major training for implementation. *Durán et al. (2023)* [57] focused on older adults with mental health needs, improving their digital literacy through the uMind mobile app.

#### Review Studies

The seven systematic reviews analyzed here, spanning research from 2016 to 2023, collectively examine digital health literacy interventions across clinical, community, and technological contexts, revealing evolving methodological approaches to evaluating eHealth and mHealth solutions, with studies assessing outcomes ranging from basic health knowledge acquisition to longitudinal behavior change in diverse populations including chronic disease patients, older adults, and socially disadvantaged groups. While demonstrating overall positive trends in health literacy improvements (particularly for underserved populations), the reviews collectively identify persistent challenges in technological accessibility, content comprehension, and long-term intervention sustainability. Jacobs et al. (2016) [12] conducted a systematic review to evaluate 12 eHealth interventions designed to improve health literacy across diverse populations, across various settings including hospitals, community centers, outpatient clinics, and worksites. The interventions, delivered through personal computers, tablets, and PDAs (Personal Digital Assistants) with web-based applications, targeted health literacy and risk behaviors related to multiple conditions including cardiovascular disease, diabetes, and colon cancer, achieved significant improvements in both health literacy levels and lifestyle behaviors compared to control groups. *Kim et al. (2017)* [58] conducted a systematic review to identify studies on online health service use by people with limited health literacy. While some positive impacts were noted, many negative results were reported related to the high reading level of online content and barriers in technology use faced by those with low health literacy. *Cheng et al. (2020)* [59] explored the role of eHealth literacy and user involvement in developing eHealth interventions targeted at socially disadvantaged groups, where one-fourth of the 51 reviewed interventions did not find statistically significant improvements in their primary outcomes, while 19 studies reported significant improvements. Long-term effectiveness also showed mixed evidence, with 8 out of 13 studies finding sustainable effects up to 12 months. *Lin et al. (2021)* [60] reviewed the effects of mHealth-based interventions on health literacy on different populations, such as patients with kidney disease, healthy adults, and adult immigrants. The interventions used diverse mHealth-based tools, including tablets, mobile phones, and PDAs, to provide nursing guidelines and information to patients, having a positive effect on health literacy, particularly among those with low education and health literacy levels. *Birati et al. (2022)* [61] conducted a scoping review to assess the literature on mHealth and telemedicine systems for women with gestational diabetes mellitus (GDM), suggesting that they can improve glycemic control and diabetes self-management in women with GDM, but there is a need for better-tailored apps that consider cultural and digital literacy. *Dong et al. (2023)* [13] reviewed the effectiveness of digital health literacy interventions for older adults, finding significant improvements in eHealth literacy, knowledge, and self-efficacy in longer-lasting interventions and those using a framework. *Verweel et al. (2023)* [15] conducted a systematic review to determine the effect of digital interventions on digital health literacy for patients with chronic diseases, finding that digital interventions generally improved digital health literacy across different chronic diseases.

See Table 4 for more details.

## 4. Discussion

Our systematic review identified multiple studies on digital interventions designed to enhance health literacy, highlighting key trends and gaps in research on health literacy interventions in the context of DHIs.

*Kim et al. (2017)* discussed the readability of online health information, with most content written above the recommended 6th-grade reading level, and suggested strategies to improve website design for low-literacy users. *Jacobs et al. (2014)* struggled with designing a tool that benefited the female breast cancer participant, calling attention to the challenge of designing informative tools for minority populations.

With regards to the application of theoretical frameworks in digital health interventions (DHIs), systematic reviews and meta-analyses (Level 1 evidence) show that theory-driven interventions yield consistently positive outcomes. *Spindler et al. (2022)*, for example, demonstrated that Self-Determination Theory (SDT) effectively increases motivation and engagement in DHIs by tapping into users’ intrinsic motivations, suggesting that theoretical rigor helps sustain user engagement over time. Quantitative studies (Levels 1–4 evidence) offer additional support; *Zoellner et al. (2020)* utilized the Theory of Planned Behavior to foster healthier attitudes toward sugar-sweetened beverage reduction, showing how behavioral models in design can enhance users’ self-efficacy and support healthier behaviors. Meanwhile, qualitative studies (Levels 4–5 evidence) like *Scull et al. (2021)* reveal that theoretical constructs, even when applied implicitly, help explain preventive health behaviors, especially in adolescents, underscoring the value of theoretically grounded design for interventions targeting behavioral change.

Several main themes emerged for discussion. User-centered design is foundational for effective DHIs. *Verweel et al. (2023)* highlighted that tools designed to fit specific user needs, such as electronic personal health records for chronic disease patients, enhanced both usability and engagement. Supporting evidence from quantitative studies (Levels 1–4 evidence) further reinforces this. *Bickmore et al. (2016)* and *Steinberg et al. (2022)* observed that user-preferred interfaces, including conversational and simplified app designs, improved health literacy, engagement, and satisfaction. When users feel supported in navigating these tools, they are more likely to achieve and maintain improved health literacy. Lower-evidence qualitative studies (Levels 4–5 evidence), such as *McKinnon et al. (2020)*, provide additional contextual understanding. These studies reveal that younger users, who may lack experience with health platforms, benefit significantly from designs incorporating visual and auditory aids, demonstrating the nuanced impact of user-centered design on engagement and usability. The findings are coherent with what is suggested by existing literature, which highlights how tailoring interventions to citizens’ needs is crucial to ensure positive attitudes and adoption [62].

Cultural competence is also essential for successful adoption and implementation of DHIs, particularly for socially disadvantaged and ethnically diverse populations. Systematic reviews and meta-analyses (Level 1 evidence) like *Cheng et al. (2020)* indicate that tailoring digital interventions to cultural and language needs significantly improves health engagement. This finding supports the view that cultural relevance fosters trust and enhances intervention efficacy. Quantitative studies (Levels 1–4 evidence), including research by *Lee et al. (2022)* and *Birati et al. (2022)*, found that language-concordant support and culturally tailored content, such as language-specific coaching, led to better health outcomes, including blood pressure control and self-management in diverse populations. These studies emphasize that integrating cultural competence into DHIs is crucial for achieving positive health outcomes. Although qualitative studies (Levels 4–5 evidence) like *Owens et al. (2020)* are less generalizable, they affirm that culturally sensitive content resonates particularly with minority groups, who value interventions reflecting their unique needs and concerns. This underscores the importance of addressing deeper cultural nuances beyond basic language adaptation.

Sustainability of health literacy gains over time is a critical goal for DHIs, as evidenced by systematic reviews and meta-analyses (Level 1 evidence). *Spindler et al. (2022)* demonstrated that sustained telerehabilitation access resulted in prolonged digital literacy improvements, indicating that follow-up and ongoing support are essential for lasting impact. Quantitative studies (Levels 1–4 evidence) show similar findings; *Scull et al. (2022)* observed that adolescents retained health literacy skills over time with periodic program engagement, suggesting that reinforcement is key for sustainable literacy gains. Qualitative studies (Levels 4–5 evidence), such as *Owens et al. (2020)*, highlight that while initial benefits are achievable, long-term adherence may be challenging without sustained user support, underlining the need for design strategies that extend beyond the intervention phase.

Active engagement strategies, including gamification and social support, are crucial for promoting sustained adherence in DHIs. Systematic reviews and meta-analyses (Level 1 evidence) show that such strategies enhance engagement, with *König et al. (2022)* reporting that gamification elements like quizzes and achievement milestones improved health literacy among adolescents. Quantitative studies (Levels 1–4 evidence) reinforce these findings; *Scull et al. (2021)* observed that social features, such as discussion boards, increased motivation and adherence, particularly among younger users. Qualitative studies (Levels 4–5 evidence), including *Jacobs et al. (2014)*, highlight the importance of personalization and user feedback mechanisms, noting that tools enabling scheduling and customized information access increase relevance and sustained use.

Accessibility and digital equity principles are essential to ensure the broad reach of DHIs, as demonstrated by systematic reviews and meta-analyses (Level 1 evidence). *Verweel et al. (2023)* found that significant health literacy improvements occurred only among populations with adequate digital access, underscoring that digital skills, connectivity, and tool access are prerequisites for equitable outcomes. Quantitative studies (Levels 1–4 evidence), like *Parker et al. (2022)*, further illustrate that socioeconomic barriers often limit engagement in lower-income populations, emphasizing the importance of low-cost or offline-compatible options. Qualitative studies (Levels 4–5 evidence) provide insight into specific access challenges; *Taha et al. (2014)* noted that older adults tend to overestimate their digital literacy, indicating the need for interventions that assess and build baseline skills to ensure equitable access.

Evaluation and outcome measurement, reliable and standardized outcome measurement is vital to assess the effectiveness of DHIs. Systematic reviews and meta-analyses (Level 1 evidence) indicate that inconsistent metrics can obscure intervention efficacy, with *Cheng et al. (2020)* emphasizing the need for rigorous outcome evaluations using standardized tools, such as eHealth Literacy Scales. Quantitative studies (Levels 1–4 evidence), such as *Scull et al. (2022)*, used validated metrics like eHEALS to track sustained improvements in media literacy, illustrating how regular assessments can identify areas needing improvement. Lower-evidence qualitative studies (Levels 4–5 evidence) add further depth, with *McKinnon et al. (2020)* noting that adolescents appreciated self-assessment features within DHIs, suggesting that integrating qualitative feedback can enhance user engagement and intervention retention.

Lastly, keeping a focus on systematic reviews and meta-analyses (Level 1 evidence), *Jacobs et al. (2016)* found that technology-based interventions, when compared to control interventions, yielded significant or promising outcomes in improving health literacy across various settings, diseases, and diverse populations. The review highlighted the feasibility of delivering eHealth interventions tailored to enhance health literacy skills for individuals with different health conditions, risk factors, and socioeconomic statuses. *Lin et al. (2021)* noted that mHealth interventions, particularly mobile applications, effectively enhance health literacy. However, the tools used to measure health literacy indicators were inconsistent, and their design often lacked specificity for assessing health literacy components. *Dong et al. (2023)* demonstrated that digital health literacy interventions positively impact the health status and management of older adults. The study emphasized the importance of practical and effective interventions to support the use of modern digital technology in managing the health of older populations.

Our results should be considered in the light of a global shift in the way healthcare is delivered. As populations age globally, the health needs of the people change, such as in response to demographic and health burden shifts in different countries. Concomitantly, technology adoption and healthcare delivery changes are happening at times when the healthcare workforce is experiencing intense shortages globally, even worsened by the migration of workers to countries with better living conditions, commonly known as the “brain drain” [63]. In this complex scenario, DHIs have the potential to change the way healthcare is delivered, shifting towards a proactive approach to healthcare, and improving population health by more precisely addressing the global burden of disease. While healthcare professionals are tackling this change by conducting innovative studies such as the ones we reviewed and investigating new forms of creation and delivery of health services [64], several international initiatives are investigating how cultural approaches guide the digital transition, particularly in a setting like healthcare. In a cross-country comparison, the Global Digital Health Partnership (GDHP) addressed the importance of digital health literacy, which emerged as the highest priority for member states. In parallel, they highlighted the importance of tailored interventions to implement digital-first healthcare systems [65].

On the policymaking side, Italy has adopted new legislation, designating a digital-first model for home care and paving the way for a new model of healthcare delivery [66].

Similarly, France [67]and Germany [68] have recently issued reimbursement models that allow the reimbursement of digital health technologies, allowing for implementing these solutions in their healthcare systems. Nonetheless, the European scenario is still fragmented on this issue, not allowing for creating a single platform for digital health solutions and instead making it difficult for innovators to deploy their solutions at an international level [69]. However, the abundance of interventions presented in our review can serve as the foundation to replicate these studies in different settings.

## 5. Limitations

Despite our adherence to best practices to ensure rigor and consistency, a degree of subjectivity is inevitable. The identified clustering of research around certain intervention types, while providing depth in these areas, may be limiting our understanding of other innovative approaches that could be particularly effective for specific populations or contexts. These patterns suggest that while digital health literacy interventions show promise, the current research landscape may be too narrowly focused, potentially hindering the development and implementation of truly innovative and inclusive solutions. The concentration of studies in specific geographical areas and intervention types, combined with the underrepresentation of certain age groups and populations, indicates a need for more diverse and comprehensive research approaches to ensure digital health literacy interventions can effectively serve all populations. One of the primary limitations of this study is its reliance on a diverse range of sources, which, although comprehensive, introduces variability in the quality and methodology of the evidence. The studies reviewed span from systematic reviews and meta-analyses (Level 1 evidence) to qualitative research (Levels 4–5 evidence), making it challenging to synthesize findings uniformly. Furthermore, the heterogeneity in study populations, interventions, and outcome measures complicate the generalizability and direct comparison of results. Many of the interventions were tailored to specific user groups, which, while necessary for cultural competence, limits the applicability to broader populations. Additionally, the study highlights the issue of digital equity, where access to technology remains a significant barrier for lower-income populations, indicating that the effectiveness of digital health interventions (DHIs) might be skewed towards those with better access to digital resources. Lastly, the evaluation and measurement of outcomes were inconsistent across studies, with some using standardized tools while others relied on less rigorous metrics, potentially affecting the validity and comparability of the results.

## 6. Conclusion

This study underscores the multifaceted nature of digital health interventions aimed at improving health literacy and engagement. It highlights that user-centered design, cultural competence, and ongoing support are crucial for the success of DHIs. Theoretical frameworks, when integrated into intervention design, enhance user engagement and behavioral change, particularly when they address intrinsic motivations and cultural nuances. The review also points to the importance of accessibility and digital equity, emphasizing that for DHIs to have a broad impact, they must address the digital divide and ensure that interventions are accessible to all, regardless of socioeconomic status or digital literacy level. Moreover, the study suggests that while short-term improvements in health literacy are achievable, sustaining these gains over time requires strategies for continuous user engagement, such as gamification or social support mechanisms. Finally, the review advocates for standardized and rigorous evaluation methods to accurately assess the effectiveness of DHIs, which would facilitate comparison across studies and inform policy and practice in digital health. The findings support the global shift towards digital-first healthcare models, with the potential to improve health outcomes, particularly in the context of an aging population and global healthcare workforce shortages. Lastly, it is essential to mention the WHO Digital Health Strategy, that serves as a potential global governance framework to establish shared objectives in digital health interventions (DHIs) and health literacy, fostering consensus on key priorities. Notably, Goal 4 of the strategy emphasizes health literacy, aligning with many of the findings in your systematic review [70].

## Data Availability

The data supporting this article is presented in Table 4, which details the final 39 papers included in this review.

## CRediT authorship contribution statement

Conceptualization: Francesco Andrea Causio, Alessandro Berionni, Luigi De Angelis, Marcello Di Pumpo, Giacomo Diedenhofen, Timothy Mackey, Fidelia Cascini

Data curation: all

Formal analysis MDP

Funding acquisition: NA

Investigation: all

Methodology: Francesco Andrea Causio, MDP

Project administration: Francesco Andrea Causio

Resources: NA

Software: NA

Supervision: Timothy Mackey, Fidelia Cascini, Caterina Rizzo, Plinio Morita

Validation: Timothy Mackey, Fidelia Cascini, Caterina Rizzo, Plinio Morita

Visualization: all

Writing – original draft: all

Writing – review & editing: all

## References

1. Berkman ND, Sheridan SL, Donahue KE, Halpern DJ, Crotty K. Low Health Literacy and Health Outcomes: An Updated Systematic Review. Ann Intern Med. 2011;155: 97–107. doi:10.7326/0003-4819-155-2-201107190-00005

2. Ninth Global Conference on Health Promotion, Shanghai 2016. [cited 22 Feb 2025]. Available: https://www.who.int/teams/health-promotion/enhanced-wellbeing/ninth-global-conference/health-literacy

3. Walters R, Leslie SJ, Polson R, Cusack T, Gorely T. Establishing the efficacy of interventions to improve health literacy and health behaviours: a systematic review. BMC Public Health. 2020;20: 1040. doi:10.1186/s12889-020-08991-0

4. Nutbeam D, McGill B, Premkumar P. Improving health literacy in community populations: a review of progress. Health Promotion International. 2018;33: 901–911. doi:10.1093/heapro/dax015

5. Classification of digital interventions, services and applications in health: a shared language to describe the uses of digital technology for health, 2nd ed. [cited 22 Feb 2025]. Available: https://www.who.int/publications/i/item/9789240081949

6. da Silva AA, Merolli M, Fini NA, Granger CL, Gustafson OD, Parry SM. Digital health interventions in adult intensive care and recovery after critical illness to promote survivorship care. Journal of the Intensive Care Society. 2025; 17511437241311105. doi:10.1177/17511437241311105

7. Senbekov M, Saliev T, Bukeyeva Z, Almabayeva A, Zhanaliyeva M, Aitenova N, et al. The Recent Progress and Applications of Digital Technologies in Healthcare: A Review. International Journal of Telemedicine and Applications. 2020;2020: 8830200. doi:10.1155/2020/8830200

8. Ran X, Chen Y, Jiang K, Shi Y. The Effect of Health Literacy Intervention on Patients with Diabetes: A Systematic Review and Meta-Analysis. IJERPH. 2022;19: 13078. doi:10.3390/ijerph192013078

9. Hollis C, Falconer CJ, Martin JL, Whittington C, Stockton S, Glazebrook C, et al. Annual Research Review: Digital health interventions for children and young people with mental health problems – a systematic and meta-review. Child Psychology Psychiatry. 2017;58: 474–503. doi:10.1111/jcpp.12663

10. Lattie EG, Adkins EC, Winquist N, Stiles-Shields C, Wafford QE, Graham AK. Digital Mental Health Interventions for Depression, Anxiety, and Enhancement of Psychological Well-Being Among College Students: Systematic Review. J Med Internet Res. 2019;21: e12869. doi:10.2196/12869

11. Scheerder A, Van Deursen A, Van Dijk J. Determinants of Internet skills, uses and outcomes. A systematic review of the second- and third-level digital divide. Telematics and Informatics. 2017;34: 1607–1624. doi:10.1016/j.tele.2017.07.007

12. Jacobs RJ, Lou JQ, Ownby RL, Caballero J. A systematic review of eHealth interventions to improve health literacy. Health Informatics J. 2016;22: 81–98. doi:10.1177/1460458214534092

13. Dong Q, Liu T, Liu R, Yang H, Liu C. Effectiveness of Digital Health Literacy Interventions in Older Adults: Single-Arm Meta-Analysis. Journal of Medical Internet Research. 2023;25: e48166. doi:10.2196/48166

14. Yeo G, Reich SM, Liaw NA, Chia EYM. The Effect of Digital Mental Health Literacy Interventions on Mental Health: Systematic Review and Meta-Analysis. J Med Internet Res. 2024;26: e51268. doi:10.2196/51268

15. Verweel L, Newman A, Michaelchuk W, Packham T, Goldstein R, Brooks D. The Effect of Digital Interventions on Related Health Literacy and Skills for Individuals Living with Chronic Diseases: A Systematic Review and Meta-Analysis. International Journal of Medical Informatics. 2023;177: 105114. doi:10.1016/j.ijmedinf.2023.105114

16. Yuen E, Winter N, Savira F, Huggins CE, Nguyen L, Cooper P, et al. Digital Health Literacy and Its Association With Sociodemographic Characteristics, Health Resource Use, and Health Outcomes: Rapid Review. Interact J Med Res. 2024;13: e46888. doi:10.2196/46888

17. Yameogo AR, Délétroz C, Sasseville M, Amil S, Da SMAR, Bodenmann P, et al. Effectiveness of Interventions to Improve Digital Health Literacy in Forced Migrant Populations: Protocol for a Mixed Methods Systematic Review. JMIR Research Protocols. 2023;12: e50798. doi:10.2196/50798

18. Causio FA, Fakhfakh M, Kaur J, Sert B, Gandolfi S, Di Pumpo M, et al. Digital Health Interventions’ Impact on Health Literacy: A Systematic Review. European Journal of Public Health. 2024;34: ckae144.1201. doi:10.1093/eurpub/ckae144.1201

19. Santana S, Brach C, Harris L, Ochiai E, Blakey C, Bevington F, et al. Updating Health Literacy for Healthy People 2030: Defining Its Importance for a New Decade in Public Health. Journal of Public Health Management and Practice. 2021;27: S258–S264. doi:10.1097/PHH.0000000000001324

20. What Is Health Literacy? | Health Literacy | CDC. [cited 12 Feb 2025]. Available: https://www.cdc.gov/health-literacy/php/about/?CDC_AAref_Val=https://www.cdc.gov/healthliteracy/learn/index.html

21. OCEBM Levels of Evidence — Centre for Evidence-Based Medicine (CEBM), University of Oxford. [cited 12 Feb 2025]. Available: https://www.cebm.ox.ac.uk/resources/levels-of-evidence/ocebm-levels-of-evidence

22. Page MJ, McKenzie JE, Bossuyt PM, Boutron I, Hoffmann TC, Mulrow CD, et al. The PRISMA 2020 statement: an updated guideline for reporting systematic reviews. Syst Rev. 2021;10: 89. doi:10.1186/s13643-021-01626-4

23. 23. Global Digital Health Monitor. [cited 22 Feb 2025]. Available: https://monitor.digitalhealthmonitor.org/map

24. Brewer LC, Fortuna KL, Jones C, Walker R, Hayes SN, Patten CA, et al. Back to the Future: Achieving Health Equity Through Health Informatics and Digital Health. JMIR Mhealth Uhealth. 2020;8: e14512. doi:10.2196/14512

25. Sukhera J. Narrative Reviews: Flexible, Rigorous, and Practical. Journal of Graduate Medical Education. 2022;14: 414–417. doi:10.4300/JGME-D-22-00480.1

26. Owens OL, Smith KN, Beer JM, Gallerani DG, McDonnell KK. A Qualitative Cultural Sensitivity Assessment of the Breathe Easier Mobile Application for Lung Cancer Survivors and Their Families. Oncology Nursing Forum. 2020;47: 331–341. doi:10.1188/20.ONF.331-341

27. Parker SM, Barr M, Stocks N, Denney-Wilson E, Zwar N, Karnon J, et al. Preventing Chronic Disease in Overweight and Obese Patients with Low Health Literacy Using eHealth and Teamwork in Primary Healthcare (HeLP-GP): A Cluster Randomised Controlled Trial. BMJ open. 2022;12: e060393. doi:10.1136/bmjopen-2021-060393

28. Steinberg JR, Yeh C, Jackson J, Saber R, Niznik CM, Leziak K, et al. Optimizing Engagement in an mHealth Intervention for Diabetes Support During Pregnancy: The Role of Baseline Patient Health and Behavioral Characteristics. Journal of Diabetes Science and Technology. 2022;16: 1466–1472. doi:10.1177/19322968211035441

29. Lemos M, Henriques AR, Lopes DG, Mendonça N, Victorino A, Costa A, et al. Usability and Utility of a Mobile App to Deliver Health-Related Content to an Older Adult Population: Pilot Noncontrolled Quasi-Experimental Study. JMIR Formative Research. 2024;8: e46151. doi:10.2196/46151

30. Chenneville T, Drake H, Gabbidon K, Rodriguez C, Hightow-Weidman L. Bijou: Engaging Young MSM in HIV Care Using a Mobile Health Strategy. Journal of the International Association of Providers of AIDS Care. 2021;20: 23259582211030805. doi:10.1177/23259582211030805

31. Reininger B, Mecca LP, Stine KM, Schultz K, Ling L, Halpern D. A Type 2 Diabetes Prevention Website for African Americans, Caucasians, and Mexican Americans: Formative Evaluation. JMIR research protocols. 2013;2: e24. doi:10.2196/resprot.2573

32. Horvath KJ, Bauermeister JA. eHealth Literacy and Intervention Tailoring Impacts the Acceptability of a HIV/STI Testing Intervention and Sexual Decision Making Among Young Gay and Bisexual Men. AIDS education and prevention: official publication of the International Society for AIDS Education. 2017;29: 14–23. doi:10.1521/aeap.2017.29.1.14

33. McKinnon KA, H Y Caldwell P, Scott KM. How Adolescent Patients Search for and Appraise Online Health Information: A Pilot Study. Journal of Paediatrics and Child Health. 2020;56: 1270–1276. doi:10.1111/jpc.14918

34. Zaim H, Keedy H, Dolce M, Chisolm D. Improving Teen Girls’ Skills for Using Electronic Health Information. Health Literacy Research and Practice. 2021;5: e26–e34. doi:10.3928/24748307-20201126-01

35. Scull T, Malik C, Morrison A, Keefe E. Promoting Sexual Health in High School: A Feasibility Study of A Web-based Media Literacy Education Program. Journal of Health Communication. 2021;26: 147–160. doi:10.1080/10810730.2021.1893868

36. Scull TM, Dodson CV, Geller JG, Reeder LC, Stump KN. A Media Literacy Education Approach to High School Sexual Health Education: Immediate Effects of Media Aware on Adolescents’ Media, Sexual Health, and Communication Outcomes. Journal of Youth and Adolescence. 2022;51: 708–723. doi:10.1007/s10964-021-01567-0

37. De Main AS, Xie B, Shiroma K, Yeh T, Davis N, Han X. Assessing the Effects of eHealth Tutorials on Older Adults’ eHealth Literacy. Journal of Applied Gerontology: The Official Journal of the Southern Gerontological Society. 2022;41: 1675–1685. doi:10.1177/07334648221088281

38. König L, Marbach-Breitrück E, Engler A, Suhr R. The Development and Evaluation of an E-Learning Course That Promotes Digital Health Literacy in School-age Children: Pre-Post Measurement Study. Journal of Medical Internet Research. 2022;24: e37523. doi:10.2196/37523

39. Conte A, Brunelli L, Moretti V, Valdi G, Guelfi MR, Masoni M, et al. Can a Validated Website Help Improve University Students’ e-Health Literacy? Annali Di Igiene: Medicina Preventiva E Di Comunita. 2023;35: 257–268. doi:10.7416/ai.2022.2542

40. Bickmore TW, Utami D, Matsuyama R, Paasche-Orlow MK. Improving Access to Online Health Information With Conversational Agents: A Randomized Controlled Experiment. Journal of Medical Internet Research. 2016;18: e1. doi:10.2196/jmir.5239

41. Moretti V, Brunelli L, Conte A, Valdi G, Guelfi MR, Masoni M, et al. A Web Tool to Help Counter the Spread of Misinformation and Fake News: Pre-Post Study Among Medical Students to Increase Digital Health Literacy. JMIR medical education. 2023;9: e38377. doi:10.2196/38377

42. Monkman H, Griffith J, Kushniruk AW. Evidence-based Heuristics for Evaluating Demands on eHealth Literacy and Usability in a Mobile Consumer Health Application. Stud Health Technol Inform. 2015;216: 358–362.

43. Chang SJ, Yang E, Lee K-E, Ryu H. Internet Health Information Education for Older Adults: A Pilot Study. Geriatric Nursing (New York, NY). 2021;42: 533–539. doi:10.1016/j.gerinurse.2020.10.002

44. Taha J, Sharit J, Czaja SJ. The Impact of Numeracy Ability and Technology Skills on Older Adults’ Performance of Health Management Tasks Using a Patient Portal. Journal of Applied Gerontology: The Official Journal of the Southern Gerontological Society. 2014;33: 416–436. doi:10.1177/0733464812447283

45. Zoellner JM, Porter KJ, You W, Estabrooks PA, Perzynski K, Ray PA, et al. The Reach and Effectiveness of SIPsmartER When Implemented by Rural Public Health Departments: A Pilot Dissemination and Implementation Trial to Reduce Sugar-Sweetened Beverages. Translational Behavioral Medicine. 2020;10: 676–684. doi:10.1093/tbm/ibz003

46. Spindler H, Dyrvig A-K, Schacksen CS, Anthonimuthu D, Frost L, Gade JD, et al. Increased Motivation for and Use of Digital Services in Heart Failure Patients Participating in a Telerehabilitation Program: A Randomized Controlled Trial. mHealth. 2022;8: 25. doi:10.21037/mhealth-21-56

47. Roh M, Won Y. Impact of Online-Delivered eHealth Literacy Intervention on eHealth Literacy and Health Behavior Outcomes among Female College Students during COVID-19. International Journal of Environmental Research and Public Health. 2023;20: 2044. doi:10.3390/ijerph20032044

48. Lee G, Chang A, Pal A, Tran T-A, Cui X, Quach T. Understanding and Addressing the Digital Health Literacy Needs of Low-Income Limited English Proficient Asian American Patients. Health Equity. 2022;6: 494–499. doi:10.1089/heq.2022.0045

49. Quialheiro A, Miranda A, Garcia M, de Carvalho AC, Costa P, Correia-Neves M, et al. Promoting Digital Proficiency and Health Literacy in Middle-aged and Older Adults Through Mobile Devices With the Workshops for Online Technological Inclusion (OITO) Project: Experimental Study. JMIR formative research. 2023;7: e41873. doi:10.2196/41873

50. Kayser L, Karnoe A, Duminski E, Jakobsen S, Terp R, Dansholm S, et al. Health Professionals’ eHealth Literacy and System Experience Before and 3 Months After the Implementation of an Electronic Health Record System: Longitudinal Study. JMIR human factors. 2022;9: e29780. doi:10.2196/29780

51. Jacobs ML, Clawson J, Mynatt ED. My Journey Compass: A Preliminary Investigation of a Mobile Tool for Cancer Patients. Proceedings of the SIGCHI Conference on Human Factors in Computing Systems. New York, NY, USA: Association for Computing Machinery; 2014. pp. 663–672. doi:10.1145/2556288.2557194

52. Jean BSt, Taylor NG, Kodama C, Subramaniam M, Casciotti D. Impacts of the HackHealth After-School Program: Motivating Youth through Personal Relevance. Proceedings of the Association for Information Science and Technology. 2015. pp. 1–11. doi:10.1002/pra2.2015.145052010032

53. Desai S, Chin J. OK Google, Let’s Learn: Using Voice User Interfaces for Informal Self-Regulated Learning of Health Topics among Younger and Older Adults. Proceedings of the 2023 CHI Conference on Human Factors in Computing Systems. Hamburg Germany: ACM; 2023. pp. 1–21. doi:10.1145/3544548.3581507

54. Pillai NM, Mohan A, Gutjahr G, Nedungadi P. Digital Literacy and Substance Abuse Awareness Using Tablets in Indigenous Settlements in Kerala. 2018 IEEE 18th International Conference on Advanced Learning Technologies (ICALT). 2018. pp. 84–86. doi:10.1109/ICALT.2018.00026

55. Wiji Utami AD, Widha Andari T, Uji Deva Satrio P, Arif S. Augmented Reality Assisted Healthy Drinking Water Products Promotion and Literacy: A Development and Evaluation. 2022 International Conference on Informatics, Multimedia, Cyber and Information System (ICIMCIS). 2022. pp. 557–562. doi:10.1109/ICIMCIS56303.2022.10017479

56. Kim H, Xie B. Health Literacy and Internet- and Mobile App-Based Health Services: A Systematic Review of the Literature. Proceedings of the Association for Information Science and Technology. 2015. pp. 1–4. doi:10.1002/pra2.2015.145052010075

57. Durán LD, Almeida AM, Figueiredo-Braga M, Lopes AC. (uMind) Mobile Application to Support Digital Literacy Interventions in Mental Health: Development and Usability Study. 2023 18th Iberian Conference on Information Systems and Technologies (CISTI). 2023. pp. 1–6. doi:10.23919/CISTI58278.2023.10211892

58. Kim H, Xie B. Health Literacy in the eHealth Era: A Systematic Review of the Literature. Patient Education and Counseling. 2017;100: 1073–1082. doi:10.1016/j.pec.2017.01.015

59. Cheng C, Beauchamp A, Elsworth GR, Osborne RH. Applying the Electronic Health Literacy Lens: Systematic Review of Electronic Health Interventions Targeted at Socially Disadvantaged Groups. Journal of Medical Internet Research. 2020;22: e18476. doi:10.2196/18476

60. Lin Y-H, Lou M-F. Effects of mHealth-based Interventions on Health Literacy and Related Factors: A Systematic Review. Journal of Nursing Management. 2021;29: 385–394. doi:10.1111/jonm.13175

61. Birati Y, Yefet E, Perlitz Y, Shehadeh N, Spitzer S. Cultural and Digital Health Literacy Appropriateness of App- and Web-Based Systems Designed for Pregnant Women With Gestational Diabetes Mellitus: Scoping Review. Journal of Medical Internet Research. 2022;24: e37844. doi:10.2196/37844

62. Causio FA, Beccia F, Kreeftenberg LL, Calabrò GE, Pastorino R, Boccia S, et al. European survey: citizens’ attitudes on personalized medicine, genetic testing and health data sharing – design and delivery. Personalized Medicine. 2024;21: 163–166. doi:10.1080/17410541.2024.2342770

63. Beccia F, Causio FA, Hoxhaj I, Huang H-Y, Wang L, Wang W, et al. Integrating China in the international consortium for personalised medicine. a position paper on healthcare professionals’ education and citizens’ empowerment in personalised medicine. BMC Med Educ. 2023;23: 438. doi:10.1186/s12909-023-04420-z

64. Causio FA, De Angelis L, Diedenhofen G, Talio A, Baglivo F. Perspectives on AI use in medicine: views of the Italian Society of Artificial Intelligence in Medicine. Journal of Preventive Medicine and Hygiene. 2024; Vol. 65 No. 2 (2024): doi:10.15167/2421-4248/JPMH2024.65.2.3261

65. Cascini F, Altamura G, Failla G, Gentili A, Puleo V, Melnyk A, et al. Approaches to priority identification in digital health in ten countries of the Global Digital Health Partnership. Front Digit Health. 2022;4: 968953. doi:10.3389/fdgth.2022.968953

66. Cascini F, Gentili A, Melnyk A, Beccia F, Causio FA, Solimene V, et al. A new digital model for the Italian Integrated Home Care: strengths, barriers, and future implications. Front Public Health. 2023;11: 1292442. doi:10.3389/fpubh.2023.1292442

67. PECAN: France’s fast-track scheme for digital health applications | ICT&health International. [cited 13 Feb 2025]. Available: https://ictandhealth.com/news/pecan-frances-fast-track-scheme-for-digital-health-applications

68. Sauermann S, Herzberg J, Burkert S, Habetha S. DiGA – A Chance for the German Healthcare System. Journal of European CME. 2022;11: 2014047. doi:10.1080/21614083.2021.2014047

69. Causio FA, Hoxhaj I, Beccia F, Marcantonio MD, Strohäker T, Cadeddu C, et al. Big data and ICT solutions in the European Union and in China: A comparative analysis of policies in personalized medicine. DIGITAL HEALTH. 2022;8: 205520762211290. doi:10.1177/20552076221129060

70. Global strategy on digital health 2020-2025. Geneva: World Health Organization; Available: https://www.who.int/publications/i/item/9789240020924

